# The role and influence of perceived experts in an anti-vaccine misinformation community

**DOI:** 10.1101/2023.07.12.23292568

**Authors:** Mallory J. Harris, Ryan Murtfeldt, Shufan Wang, Erin A. Mordecai, Jevin D. West

## Abstract

The role of perceived experts (i.e., medical professionals and biomedical scientists) as potential anti-vaccine influencers has not been characterized systematically. We describe the prevalence and importance of anti-vaccine perceived experts by constructing a coengagement network based on a Twitter data set containing over 4.2 million posts from April 2021. The coengagement network primarily broke into two large communities that differed in their stance toward COVID-19 vaccines, and misinformation was predominantly shared by the anti-vaccine community. Perceived experts had a sizable presence within the anti-vaccine community and shared academic sources at higher rates compared to others in that community. Perceived experts occupied important network positions as central anti-vaccine nodes and bridges between the anti- and pro-vaccine communities. Perceived experts received significantly more engagements than other individuals within the anti- and pro-vaccine communities and there was no significant difference in the influence boost for perceived experts between the two communities. Interventions designed to reduce the impact of perceived experts who spread anti-vaccine misinformation may be warranted.

## 2 Introduction

Vaccine refusal poses a major threat to public health, and has been a particular concern during the COVID-19 pandemic [Islam et al., 2020, Tangcharoensathien et al., 2020, Bonnevie et al., 2021, Carpiano et al., 2023]. An estimated 232,000 vaccine-preventable COVID-19 deaths occurred in unvaccinated adults in the United States across a fifteen-month period (May 2021 - September 2022) [Jia et al., 2023]. Exposure to misinformation (i.e., false or misleading claims) may reduce vaccine uptake, increasing individual risk of morbidity and mortality and potentially lead to disease outbreaks [Loomba et al., 2021, Wilson and Wiysonge, 2020]. The Internet, particularly social media, is an important source of both vaccine information and misinformation [Jones et al., 2012, Kata, 2010, Wawrzuta et al., 2021, Islam et al., 2020]. Social media surveillance has been proposed as a strategy to assess public opinion about vaccination and to study patterns in vaccine decision-making that may inform interventions [Cascini et al., 2022, Tavoschi et al., 2020, Boucher et al., 2021, Griffith et al., 2021, Tangcharoensathien et al., 2020]. For example, online social networks often contain *influencers*, users who play outsized roles in information propagation and receive significantly more engagement with their content than other users [Peng et al., 2017, Bakshy et al., 2011, Cha et al., 2010, Goyal et al., 2008, DeVerna et al., 2022]. Once identified, influencers may be targeted to optimize rapid dissemination of information (e.g., for advertising, public service announcements, or fact-checking purposes) [Peng et al., 2017, Bakshy et al., 2011, Nguyen et al., 2013, Candogan and Drakopoulos, 2017] or to reduce the propagation of harmful content with minimal intervention [Smith et al., 2021, DeVerna et al., 2022, Manoel Horta Ribeiro et al., 2018].

Information consumers often use markers of credibility to assess different sources [Acerbi, 2016, Sundar, 2007]. Specifically, *prestige bias* describes a heuristic where one preferentially learns from individuals who present signals associated with higher status (e.g., educational and professional credentials) [Jiménez and Mesoudi, 2019, Jucks and Thon, 2017, Berl et al., 2021]. Importantly, prestige-biased learning relies on signifiers of expertise that may or may not be accurate or correspond with actual competence in a given domain [Acerbi, 2016, Brand et al., 2020]. Therefore we will refer to *perceived experts* to denote individuals and organizations that signal biomedical expertise in their social media profiles, but note that credentials may be misrepresented or misunderstood. In particular, we focus on the understudied role of perceived experts as potential anti-vaccine influencers who accrue influence through prestige bias [Cascini et al., 2022, Tangcharoensathien et al., 2020]. Medical professionals, biomedical scientists, and organizations are trusted sources of medical information who may be especially effective at persuading people to get vaccinated and correcting misconceptions about disease and vaccines [Amazeen and Krishna, 2020, Vraga and Bode, 2017, Freed et al., 2011, Gesser-Edelsburg et al., 2018, Jucks and Thon, 2017], suggesting that prestige bias may apply to vaccination decisions, including for COVID-19 vaccines [Lopes and 2021, 2021, Reiter et al., 2020, Berenson et al., 2021].

*Despite the large body of research on perceived experts who recommend vaccination, the prevalence and influence of perceived experts acting in the opposite role, as disseminators of false and misleading claims about health has not been studied directly*. Prior work on anti-vaccine influencers suggested a category analogous to our definition of perceived experts and provided notable examples [Smith, 2017, Van Schalkwyk, 2019, The Virality Project, 2022, Carpiano et al., 2023]. For example, former physician Andrew Wakefield and other perceived experts promulgated the myth that the measles, mumps, and rubella (MMR) vaccine causes autism [Kata, 2010], perceived experts appeared in the viral Plandemic conspiracy documentary and other anti-vaccine films [Prasad, 2022, Bradshaw et al., 2020, 2022], and six of the twelve anti-vaccine influencers identified as part of the “Disinformation Dozen” responsible for a majority of anti-vaccine content on Facebook and Twitter included medical credentials in their social media profiles [Center for Countering Digital Hate, 2021]. Anti-vaccine users comprised a considerable proportion of apparent medical professionals (a subset of perceived experts excluding scientific researchers) sampled based on use of a particular hashtag (#DoctorsSpeakUp) [Bradshaw, 2022] or inclusion of certain keywords in their profiles [Ahamed et al., 2022, Kahveci et al., 2022]. The number and population share of perceived experts across the community of individuals on the microblogging website Twitter opposing COVID-19 vaccines has not been assessed systematically, an important step toward understanding the scale of this set of potential anti-vaccine influencers and one of the goals of this paper.

In addition to signaling expertise in their profiles, perceived experts may behave like biomedical experts by making scientific arguments and sharing scientific links, but also propagate misinformation by sharing unreliable sources. Anti-vaccine documentaries frequently utilize medical imagery and emphasize the scientific authority of perceived experts who appear in the films [Prasad, 2022, Bradshaw et al., 2020, Hughes et al., 2021]. Although vaccine opponents reject scientific consensus, many still value the brand of science and engage with peer reviewed literature [Koltai and Fleischmann, 2017]. Scientific articles are routinely shared by Twitter users who oppose vaccines and other public health measures (e.g., masks), but sources may be presented in a highly selective or misleading manner [van Schalkwyk et al., 2020, Van Schalkwyk, 2019, Abhari et al., 2023, Lee et al., 2021, Beers et al., 2022, Koltai and Fleischmann, 2017]. At the same time, misinformation claims from sources that often fail fact checks (i.e., low-quality sources) are pervasive within anti-vaccine communities, where they may exacerbate vaccine hesitancy [Loomba et al., 2021, Sharma et al., 2021, DeVerna et al., 2022, Muric et al., 2021]. Here, we investigate how much perceived experts in the anti-vaccine community share both misinformation and scientific sources compared to other users to understand the types of evidence perceived experts use to support their arguments.

After describing the types of information that perceived experts share, we evaluate their ability to reach large audiences who help spread their messages. Various network centrality metrics have been developed to describe the importance of a given node (in this case, user) to information flow based on connections to other nodes. For example, *degree centrality* is the count of edges (connections between nodes) linking a given node to other nodes, *betweenness centrality* is the number of times a given node lies on the shortest path between two other nodes, and *eigenvector centrality* scores nodes recursively based on the centrality of the nodes to which they are connected (Supplemental Table 1). Centrality metrics are commonly used to rank the importance of different users within a social media and help identify influential users [Cha et al., 2010, Hagen et al., 2018, Simmie et al., 2014, Riquelme and Gonźalez-Cantergiani, 2016, Gilbert and Paulin, 2015]. Pro-vaccine perceived experts were highly central in other Twitter networks discussing vaccines, but no prior analysis has focused on the the centrality of perceived experts in the anti-vaccine community [Hagen et al., 2022, Sanawi et al., 2017]. In addition to a node’s centrality to the whole network, its ability to span opposing communities may be particularly significant [Beers et al., 2023]. “Cognitive bridges,” or users who share content of interest to both anti-vaccine and pro-vaccine communities may be particularly important due to their potential to connect vaccine skeptics with accurate information or to reduce vaccine confidence in pro-vaccine audiences [van Schalkwyk et al., 2020]. If perceived experts occupy central and bridging network positions, they may be well-positioned to share their opinions with other users and share (mis)information about vaccines.

In general, perceived expertise may increase user influence within anti-vaccine communities. Although vaccine opponents express distrust in scientific institutions and the medical community writ large, they simultaneously embrace perceived experts who oppose the scientific consensus as heroes and trusted sources [Kata, 2010, Prasad, 2022, Hughes et al., 2021, Bradshaw et al., 2022]. Medical misinformation claims attributed to perceived experts were some of the most popular and durable topics within misinformation communities on Twitter during the COVID-19 pandemic [Boucher et al., 2021, Haupt et al., 2021]. In fact, compared to individuals who agree with the scientific consensus, individuals who hold counter-consensus positions may actually be more likely to engage with perceived experts that align with their stances [Beers et al., 2022, Faasse et al., 2016, Wood, 2018]. This expectation is based on experimental work on *source-message incongruence*, which suggests that messages are more persuasive when they come from a surprising source [Manes, 2019, Wood and Eagly, 1980]. This phenomenon may extend to the case where perceived experts depart from the expected position of supporting vaccination. In an experiment where participants were presented with claims from different sources about a fictional vaccine, messages from doctors opposing vaccination were especially influential and were transmitted more effectively than pro-vaccine messages from doctors [Jiménez et al., 2018]. However, this effect has not been tested for actual perceived experts commenting on real vaccines. We posit that perceived experts who spread misinformation may be disproportionately amplified within the Twitter network and have a greater influence on opinion compared to those who advance consensus positions and encourage vaccination [Efstratiou and Caulfield, 2021]. By quantifying the relative impact of perceived experts within the anti-vaccine community compared to other individuals, we will establish whether they represent a particularly influential group that should be specifically considered in interventions to encourage vaccine uptake.

For this study, we collected over 4.2 million unique posts to Twitter containing keywords about COVID-19 vaccines during April 2021 (listed in Supplemental File 1). Users with shared audiences (i.e., those whose posts were shared at least twice by at least ten of the same users) were linked to form a coengagement network Figure 1, which is analogous to co-citation networks in bibliographic analyses that connect sources that are commonly referenced together [Beers et al., 2023]. This method groups users based on how their tweets are received instead of how they engage with other users (as would be the case if edges were based on whether users retweeted, mentioned, or followed each other) and filters the network to users who were retweeted by people who also retweeted other people in the network (i.e., users who have an audience). Profile names and descriptions for all accounts in the coengagement network were reviewed to identify individual users with English profiles and classify those with professional or academic biomedical credentials in their profiles as perceived experts. The network was separated into two main communities using the Infomap community detection algorithm, which identifies densely interconnected groups of nodes. Three coders assessed a sample of popular tweets from both communities and determined that users in one generally expressed a negative stance toward COVID-19 vaccines while users in the other were primarily positive, leading us to label the communities as antiand pro-vaccine respectively.

**Figure 1:**
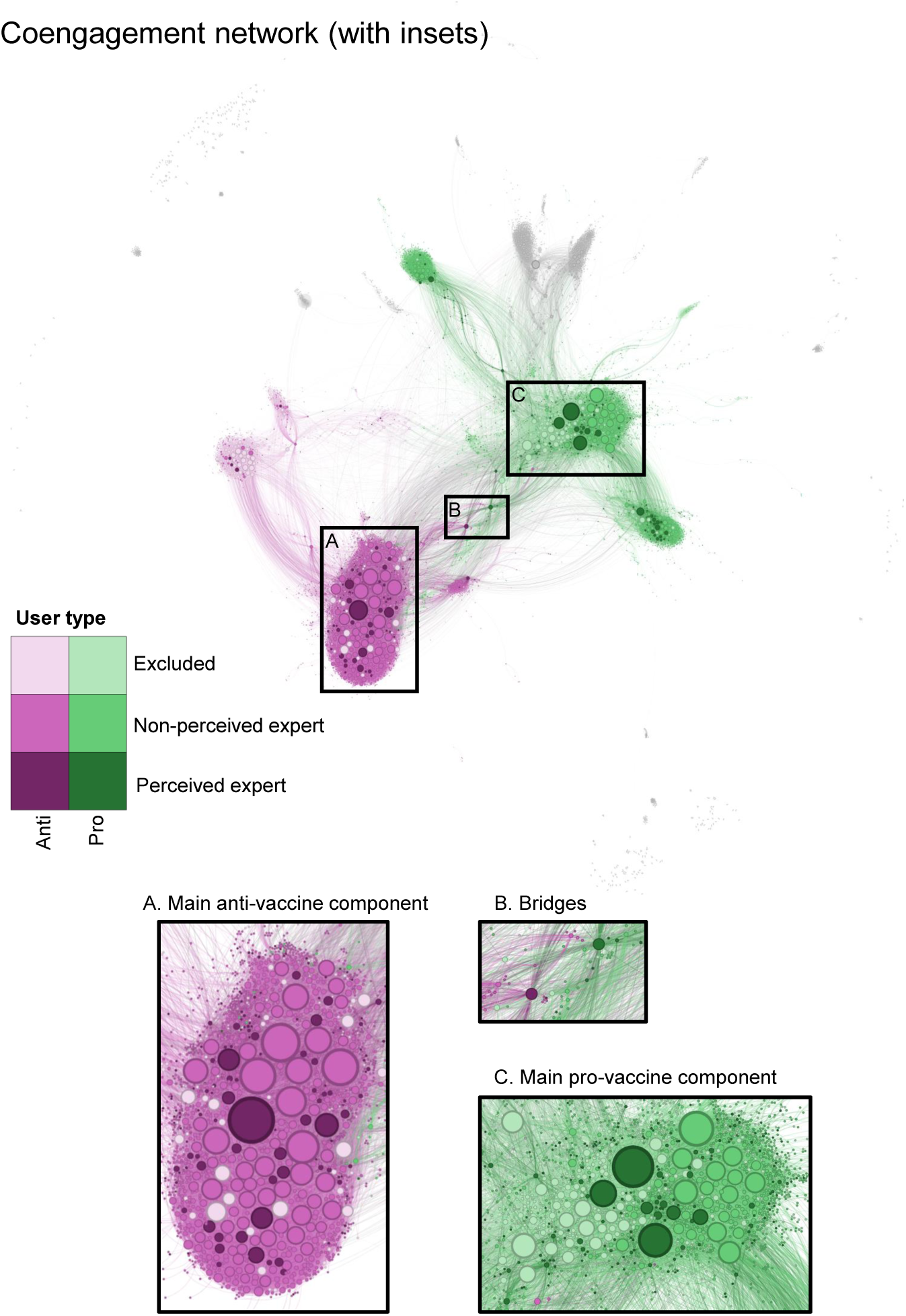
The coengagement network of users tweeting about COVID-19 vaccines is divided into two large communities. Users are represented as circles and scaled by degree centrality. Edges connect users that were retweeted at least twice by at least ten of the same users. Nodes in the two largest communities detected using the Infomap algorithm are colored in pink (anti-vaccine) and green (pro-vaccine). Shades indicate account type: excluded from analyses (light); individual perceiv7ed non-expert (medium); and perceived expert (dark). Nodes outside of the two largest communities are gray. Insets provide more detailed views of: (A) the main component of the anti-vaccine community, (B) bridges between the pro- and anti-vaccine communities, and (C) the main component of the pro-vaccine community. Each edge is colored based on the color of one of the two nodes it connects, randomly selected. A higher-resolution image without annotations is available as Supplemental Figure 6.

Our analysis is divided into two parts, where the first focuses on describing the community of perceived experts and the second evaluates their influence relative to other individuals. First, we characterized the prevalence of perceived experts and roles they play in conversations about COVID-19 vaccines, asking: (1) How many perceived experts are there in the anti- and pro-vaccine communities? and (2) How often do perceived experts in the anti-vaccine community share misinformation and scientific sources relative to other users? We first calculated the number and proportion of perceived experts in the anti- and pro-vaccine communities. To assess the types of evidence shared in the anti- and pro-vaccine communities depending on perceived expertise, we determined the proportion of links shared from domain names classified as low-quality sources by Media Bias/Fact Check or as academic by databases of research publications [Newbold et al., 2022] and pre-print servers [Kirkham et al., 2020].

Second, we tested whether perceived experts are particularly important in discussion around COVID-19 vaccines, asking: (3) Do perceived experts occupy key positions within the coengagement network (i.e., as central and bridging nodes)? and (4) Are perceived experts more influential than other individual users? First, we tested whether perceived experts were disproportionately part of the group of highly central users in the anti-vaccine and pro-vaccine community based on degree, betweenness, and eigenvector centrality Table 1. We also tested whether perceived experts were overrepresented among the users with the greatest community bridging scores relative to their share of the population. Second, we conducted propensity score matching to assess whether, on average, perceived experts received more engagements (i.e., likes and retweets) and had greater network centrality compared to other users with similar profile characteristics and posting behaviors who were not perceived experts. We conducted this analysis for both communities separately, hypothesizing that: perceived experts experienced an influence boost in both the anti-vaccine community (H1) and the pro-vaccine community (H2). We then compared the size of this effect between the two communities, hypothesizing that perceived experts experienced a larger influence boost within the anti-vaccine community due to source-message incongruence (H3).

## 3 Results

### 3.1 Perceived experts are present throughout the coengagement network, including in the anti-vaccine community

The coengagemement network consisted of 7,720 accounts (nodes) linked by 72,034 edges (Figure 1). 5,171 of those accounts had English language profiles and appeared to correspond to individual users. 24 of the individual users with English profiles added or removed expertise cues in their profiles over the course of the study and were thus excluded from the analysis. Of the remaining 5,147 individual users, 13.1% (678 users) were perceived experts. Perceived experts rarely provided cues of expertise in their name alone. Instead, almost all users indicated expertise in their description, with approximately half of users including expertise cues in both their name and description (Supplemental Figure 7). There was substantial agreement between coders on whether a user was in an excluded category, a perceived non-expert, or a perceived expert (*κ* = 0.715).

The two largest communities contained 79.6% of total accounts and 66.1% of English-language individual accounts in the network. Stance toward COVID-19 vaccines by users with the greatest degree centrality was relatively consistent, meaning that very few users posted a combination of tweets that were positive and negative in stance, although most users posted some neutral tweets as well (Supplemental Figure 8). Further, stance was generally shared within communities; popular tweets by central users in the anti- and pro-vaccine communities almost exclusively expressed negative and positive stances, respectively, or neutral stances, with a few exceptions. Although there was moderate inter-rater reliability (*κ* = 0.567) between coders on whether individual tweets were negative, positive or neutral about vaccines, there was high agreement on the overall stance of each user (*κ* = 0.805 for whether a given user posted more anti-vaccine tweets, pro-vaccine tweets, or equal numbers of either).

The pro-vaccine community was larger than the anti-vaccine community (3,443 and 2,704 users respectively). The anti-vaccine community was made up of a large, densely connected component split into two communities and linked to two additional medium-sized subcommunities (Figure 1A), which were dominated by users tweeting in languages other than English (Italian and French) (see Supplemental Figure 10 and Supplemental Table 4 for visualization and further discussion of subcommunities). The pro-vaccine community similarly consisted of one large component that was connected to the next largest community, which was focused on vaccination in India, and two medium-sized subcommunities (Figure 1C). The largest component of the pro-vaccine community was divided into two subcommunities, one of which was dominated by non-individual accounts. Perceived experts were present in both communities, but constituted a larger share of individual users in the pro-vaccine community (17.2 % or 386 accounts) compared to the anti-vaccine community (9.8 % or 185 accounts) (Figure 3). The two communities were connected by a few bridging nodes, including some perceived experts (Figure 1B).

### 3.2 Perceived experts in the anti-vaccine community share both low quality and academic sources

We found marked differences in sharing of low quality and academic sources depending on community and perceived expertise Figure 2. For both types of sources, we calculated a user-level metric (the proportion of users who shared at least one link of a given type; Figure 2B, D) and a link-level metric (the proportion of all links shared during the study period that were of a given type; Figure 2A, C). Perceived experts posted more frequently, and included links in a greater proportion of their posts (Supplemental Figure 9)

**Figure 2:**
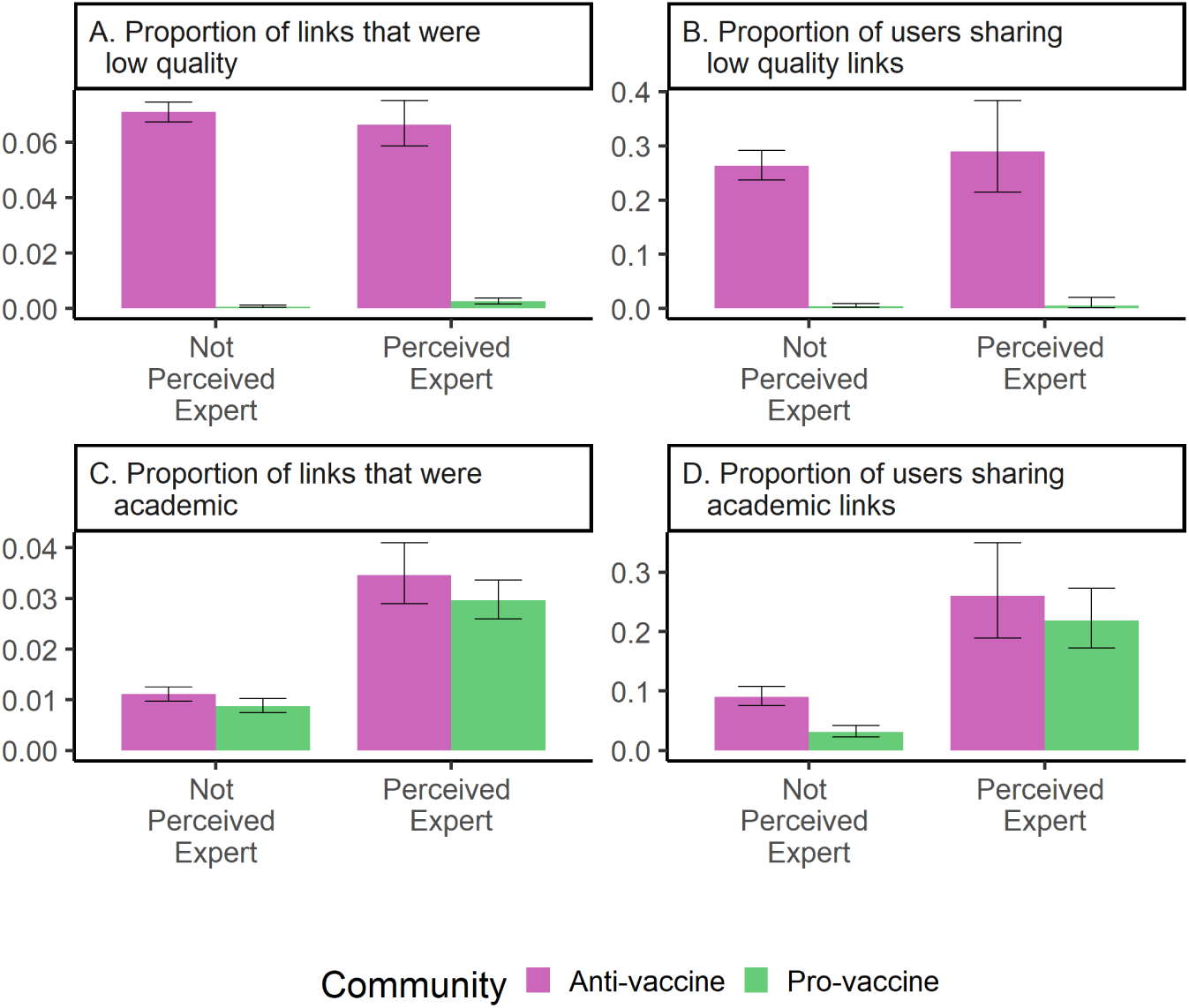
Users in the anti-vaccine community share low quality sources and perceived experts share academic research. Each panel compares a different metric of link-sharing by perceived experts and perceived non-experts in the anti(pink) and pro-(green) vaccine communities. The metrics are: (A) proportion of checked links that were from low quality sources, (B) proportion of users that shared at least one low quality source, (C) proportion of checked links that were from academic research sources, and (D) proportion of users that shared at least one academic research source. 95% binomial proportion confidence intervals are indicated by black error bars.

Compared to the pro-vaccine community, perceived experts and perceived non-experts in the anti-vaccine community shared low quality sources at significantly greater rates (*p <* 0.001 for proportion of links and proportion of users). Low quality sources were almost exclusively shared in the anti-vaccine community, although low quality sources generally comprised a relatively small proportion of assessed links (Figure 2A, B). Many users in the anti-vaccine community shared at least one low quality link (29% of perceived experts and 26% of perceived non-experts) compared to fewer than 0.5% of users in the pro-vaccine community (two perceived experts and seven perceived non-experts) (Figure 2B).

In both communities, perceived experts shared academic research links at a significantly greater rate compared to other individual users (Figure 2C, D) (*p <* 0.001 for proportion of links and proportion of users). There was greater academic link-sharing by perceived non-experts in the anti-vaccine community compared to the pro-vaccine community (*p* = 0.011 and *p <* 0.001 for proportion of links and proportion of users respectively).

### 3.3 Perceived experts are overrepresented as central and bridging users

Although perceived experts represented a relatively small share of the individual users in the coengagement network, they disproportionately occupied important positions in the network as central and bridging nodes (Figure 3, Figure 4). Perceived experts were overrepresented among users with the greatest betweenness, degree, and eigenvector centrality in both communities, although this effect was not always statistically significant for the pro-vaccine community (Figure 3).

Perceived experts in both communities were highly overrepresented among the five hundred users with the greatest betweenness centrality (*p <* 0.001 for both communities) and overrepresented among the fifty users with the greatest betweenness centrality by a factor of about two (*p* = 0.014 and *p <* 0.001 for the anti- and pro-vaccine communities respectively) (Figure 3).

Ranking on degree centrality and eigenvector centrality, perceived experts were more strongly overrepresented in the anti-vaccine community compared to the pro-vaccine community (Figure 3). Perceived experts in the anti-vaccine community were about two times more prevalent in the group of central users (20% of the fifty top users ranked on both metrics) compared to their share of the population, while perceived experts in the pro-vaccine community were overrepresented by a factor of approximately 1.6. By both metrics, perceived experts in the anti-vaccine community were significantly overrepresented in the 500 most central users (*p* = 0.001 and *p <* 0.001 for degree and eigenvector centrality respectively) and the fifty most central users (*p* = 0.014 for both degree and eigenvector centrality). In the pro-vaccine community, perceived experts were significantly overrepresented in the 50 most central users (*p* = 0.032 for both degree and eigenvector centrality) but not in the 500 most central users (*p* = 0.1038 and *p* = 0.082). It is unsurprising that the results for eigenvector and degree centrality were similar given that the PageRank algorithm used to compute eigenvector centrality is proportional to degree centrality on an undirected graph [Grolmusz, 2015].

Perceived experts were significantly overrepresented in the group of the 500 and ten most important bridges between the pro- and anti-vaccine communities (*p <* 0.001, *p* = 0.001 respectively), but not significantly overrepresented in the fifty most important bridges (*p* = 0.09). Figure 4. About 20% of the top 500 and top fifty users ranked by community bridging scores were perceived experts, and five of the 10 users with the greatest community bridging scores were perceived experts (Figure 4, Figure 1B). There was little variation in which users were highly ranked between different network metrics (Supplemental Figure 12, Supplemental Figure 13, Supplemental Figure 14, Supplemental Figure 15).

**Figure 3:**
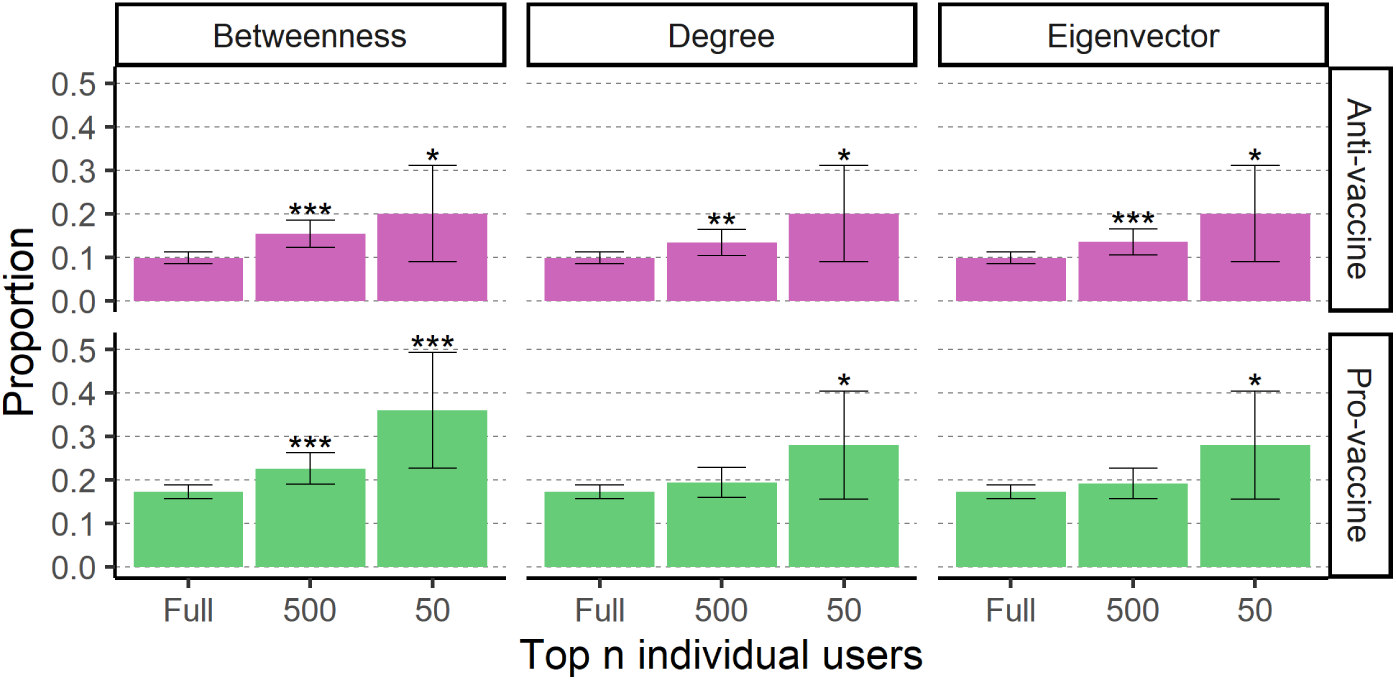
Perceived experts are overrepresented as the most central users in the pro-vaccine (green) and anti-vaccine (pink) communities. Plots are arranged in a grid where each row corresponds to users in one of the two largest communities: the anti- and pro-vaccine communities (top and bottom respectively). Each column corresponds to a different centrality metric: betweenness centrality (left), degree centrality (middle), and eigenvector centrality (right). For each plot, we subset the network to the *n* nodes with the greatest values for a given centrality metric, where *n* is full population size (all individual users in the community), 500, or 50 (x-axis). Bar height indicates the proportion of users in each subset that are perceived experts and error bars give 95% binomial proportion confidence intervals. Stars above the error bars indicate whether perceived experts are significantly overrepresented within a given sample of central users (one star indicates *p <* 0.05, two stars indicate *p <* 0.01, three stars indicate *p <* 0.001).

**Figure 4:**
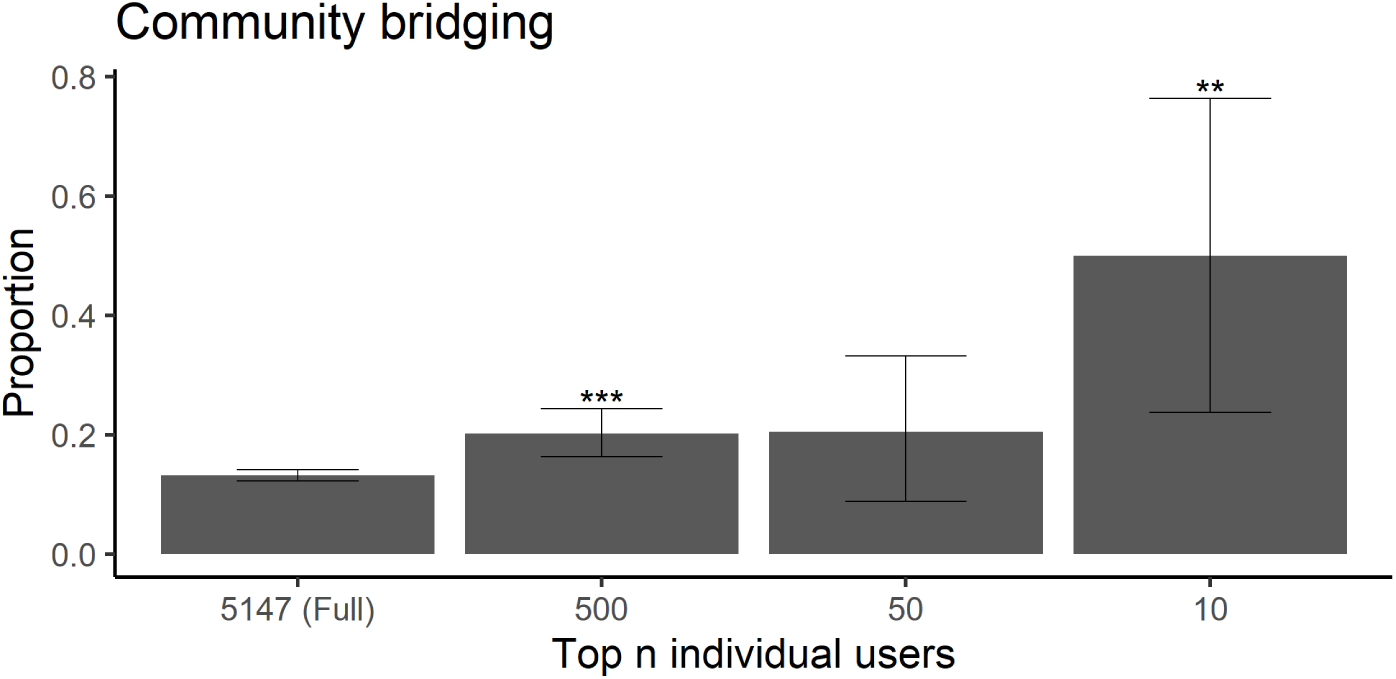
Perceived experts disproportionately act as key bridges between the pro-vaccine and anti-vaccine communities. Bar height indicates the proportion of users in each population sample that are perceived experts and error bars give 95% confidence intervals for the proportions. The x-axis indicates the size of each subset (*n*), corresponding to the full population of 5,147 (all individual users as a basis of comparison), and the 500, 50, or 10 users with the greatest community bridging score (x-axis). Stars above the error bars indicate whether perceived experts are significantly overrepresented within a given sample of bridging users (one star indicates *p <* 0.05, two stars indicate *p <* 0.01, three stars indicate *p <* 0.001).

### 3.4 Perceived experts are more influential than other individuals in the anti- and pro- vaccine communities

Using propensity score matching, we achieved excellent balance across matching covariates within the anti-vaccine community (Supplemental Figure 16) and good matching within the pro-vaccine community for all variables except for the percent of posts with links (Supplemental Figure 17 and Supplemental Figure 18). There were minimal differences in the results of propensity score matching with and without matching on subcommunities (compare Figure 5 and Supplemental Figure 19; Supplemental Table 6, Supplemental Table 7, and Supplemental Table 8). Using these propensity-matched pairs for comparison, on average, perceived experts received more engagements (likes and retweets) on their posts, but were not more central than other individual users (Figure 5, Supplemental Figure 19, Supplemental Table 6, Supplemental Table 7, Supplemental Table 8).

**Figure 5:**
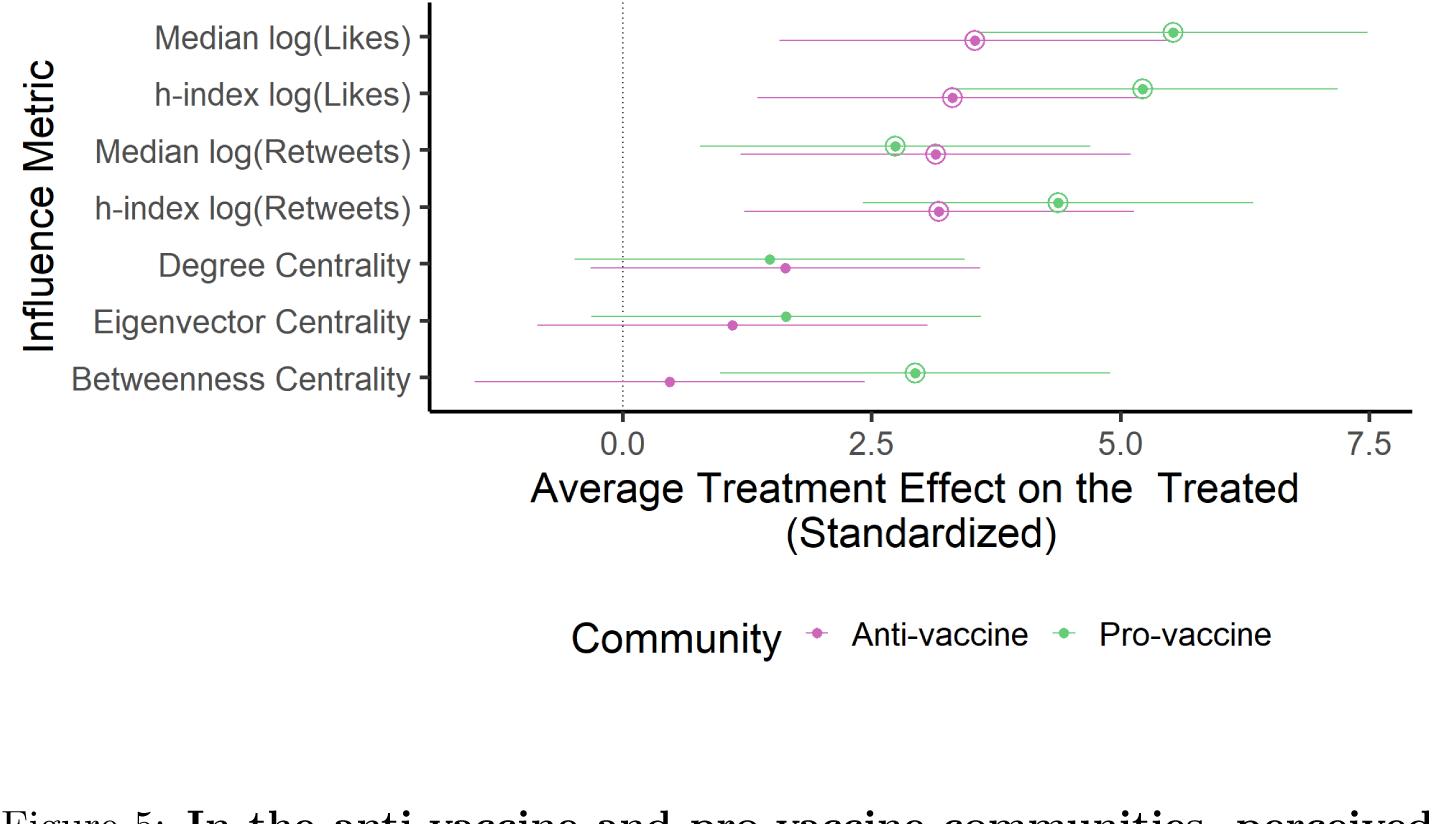
In the anti-vaccine and pro-vaccine communities, perceived experts receive greater engagements compared to other users. For each influence metric (y-axis, Supplemental Table 3), we plot the standardized average treatment effect on the treated (ATT) as a point and corresponding 95% confidence interval. Values are shown for the matching analysis directly comparing the anti-vaccine (pink) and pro-vaccine (green) communities (corresponding to Supplemental Table 8). Positive values (to the right of the vertical line) indicate an influence boost for perceived experts. Instances where the effects were significantly greater than zero (*p <* 0.05) are indicated with an additional circle around the point estimate.

Perceived experts in the anti-vaccine community received 20% (95% CI: 3 - 43%) more retweets and 37% (95% CI: 9 - 63%) more likes on their median post than would be expected if they were not perceived experts (Figure 5, Supple-mental Figure 19, Supplemental Table 6). We also found a significant effect of perceived expertise when using an alternative metric, the h-index, to assess engagements across tweets posted during the time period (ATT for natural logged retweets: 1.90 (95% CI: 0.36-3.44); ATT for natural logged likes: 3.43 (95% CI: 1.11-5.76)) (Figure 5, Supplemental Figure 19, Supplemental Table 6). In contrast to these measures of engagement, perceived expertise did not significantly affect centrality (betweenness, degree, and eigenvector) within the anti-vaccine community *on average* (Figure 5, Supplemental Table 8, Supplemental Table 6), although we found in the previous section that perceived experts were overrepresented in the *tail* of the distribution as highly central users compared to the full (unmatched) set of perceived non-experts (see subsection 7.8 for further discussion reconciling these two results).

Perceived experts in the pro-vaccine community also had a positive ATT across engagement metrics and additionally had a significantly positive ATT for betweenness centrality (Figure 5, Supplemental Figure 19, Supplemental Table 7). Although perceived experts in the pro-vaccine community tended to have a greater ATT across all influence metrics than those in the anti-vaccine community, the differences in ATT between groupswere not statistically significant for any influence metric (Supplemental Figure 20, Figure 5, Supplemental Table 9). Matching results were robust to matching parameters and exclusion of different matching covariates (Supplemental Figure 22, Supplemental Figure 23, and Supplemental Figure 24).

## 4 Discussion

The anti-vaccine community contains its own set of perceived experts. These perceived experts represent 9.8% of individual users within the anti-vaccine community, comprising a substantial group that extends beyond the handful of high-profile anti-vaccine influencers with biomedical credentials who have been noted anecdotally [Smith, 2017, The Virality Project, 2022]. Although surveys have found broad support for COVID-19 vaccination among medical providers [Callaghan et al., 2022, Bartoš et al., 2022], 28.9% of perceived experts in the two largest communities of the coengagement network were part of the anti-vaccine community, a proportion similar to those reported by other studies of COVID19 vaccine attitudes expressed by medical professionals on Twitter [Ahamed et al., 2022, Kahveci et al., 2022, Bradshaw, 2022]. Anti-vaccine perceived experts are therefore overrepresented on Twitter compared to the share of actual biomedical experts who oppose Covid-19 vaccines, which may lead observers to underestimate the scientific consensus in favor of COVID-19 vaccination, in turn reducing vaccine uptake [Efstratiou and Caulfield, 2021, Bartoš et al., 2022, Motta et al., 2023].

Within the anti-vaccine community, perceived experts may combine misinformation with claims that appear scientific. Low quality sources in the coengagement network were overwhelmingly shared by the anti-vaccine community, and perceived experts shared low quality sources at similar rates compared to other individuals (Figure 2), suggesting that they directly contribute to the widespread misinformation in the anti-vaccine community noted by other studies [Memon and Carley, 2020, Muric et al., 2021, Jamison et al., 2020]. Perceived experts, including those in the anti-vaccine community, performed expertise by sharing and commenting on academic articles at much higher rates compared to other individual users (Figure 2C, D). Misinformation claims containing arguments that appear scientific may be particularly effective at reducing vaccine intent [Loomba et al., 2021], suggesting that perceived experts may be responsible for some of the most compelling anti-vaccine claims.

Perceived experts may also be well-poised to spread their claims online, as they disproportionately occupied key network positions between anti- and pro-vaccine communities and within the anti-vaccine community (Figure 3, Figure 4). Across various centrality metrics, perceived experts were overrepresented in the group of highly central users Figure 3, meaning that they reached (and had posts shared by) large and unique audiences. Perceived experts were half of the ten individuals users with the greatest community bridging scores, meaning that they were shared by audiences for users in both the anti- and pro-vaccine communities. Within this set of five perceived experts, four made highly technical arguments in favor of vaccines and corrected potential misunderstandings related to the frequency of vaccine-linked adverse events and to the severity of breakthrough infections reported at the time. Such users could play an important role in changing vaccine stance, although the two communities may share distinct subsets of their tweets for substantially different reasons [Van Schalkwyk, 2019, Beers et al., 2023]. For example, anti-vaccine audiences may retweet reports of adverse events out of concern that vaccines are unsafe while pro-vaccine audiences may share the same content to emphasize their rarity.

Our hypothesis that perceived experts are, on average, more influential than other users in the anti-vaccine community was supported by the finding that they received more engagement (i.e., likes and retweets) on their vaccine-related posts than similar users without credentials in their profiles Figure 5. We also found evidence of this effect in the pro-vaccine community. Perceived experts were not significantly more central on average than a matched set of perceived non-experts (except in the case of perceived experts in the pro-vaccine community who had greater betweenness centrality than perceived non-experts) Figure 5. These findings may be explained by the observations that matching covariates (e.g., follower count, post frequency) contribute importantly to centrality (Supplemental Figure 21) and that overrepresentation of perceived experts in the set of highly central users (i.e., the tail of the distribution) may not be sufficient to significantly increase the mean centrality of perceived experts compared to perceived non-experts Figure 3. There was no significant difference in the influence boost for perceived experts between the anti- and pro-vaccine communities, contradicting our hypothesis that perceived experts hold a greater advantage within the anti-vaccine community. However, this finding suggests that anti-vaccine audiences do value expert opinion, at least when it confirms their own stances, a result aligned with other studies that have found that scientists and medical professionals are popular sources amongst vaccine opponents [Koltai and Fleischmann, 2017, Kata, 2010, Prasad, 2022, Hughes et al., 2021, Bradshaw et al., 2022, Boucher et al., 2021, Haupt et al., 2021].

In sum, this works goes beyond high-profile examples of anti-vaccine perceived experts to systematically characterize the sizable population of anti-vaccine perceived experts who have a significant impact in the the Twitter conversation about COVID-19 vaccines. Our findings have implications for interventions focused on education of medical professionals and the general public. Educational interventions that encourage trust in science could backfire if individuals defer to anti-vaccine perceived experts who share low quality sources [O’Brien et al., 2021]. Instead, education efforts should focus on teaching the public how to evaluate source credibility to counter the potentially fallacious heuristic of deferring to individual perceived experts [O’Brien et al., 2021, Osborne and Pimentel, 2022, Sundar, 2007]. Although the sample of perceived experts in this study is not representative of the broader community of experts, surveys have found that a non-negligible minority of medical students and health professionals are vaccine hesitant and believe false claims about vaccine safety [Lucia et al., 2021, Mose et al., 2022, Callaghan et al., 2022, Le Marechal et al., 2018]. Given that perceived experts are particularly influential in vaccine conversations (Figure 3, Figure 4, Figure 5) and that healthcare providers with more knowledge about vaccines are more willing to recommend vaccination, efforts to educate healthcare professionals and bioscientists on vaccination and to overcome misinformation within this community may help to improve vaccine uptake [Paterson et al., 2016].

In addition to helping people evaluate vaccine information, interventions may focus on improving information quality by focusing on communications, social media platform design, and expert community self-governance. From a communication standpoint, greater engagement by perceived experts recommending COVID-19 vaccines and debunking medical misinformation may help to correct public misunderstandings about expert consensus based on the overrepresentation of anti-vaccine perceived experts on social media [Bartoš et al., 2022]. However, perceived experts already constituted one fifth of individuals in the pro-vaccine community according to our analysis [Hernandez et al., 2021, Gallagher et al., 2021, Herńandez-Garćıa et al., 2021] and it is not clear whether individual pro-vaccine communicators will be especially persuasive to people who are already engaging with perceived experts who oppose vaccines. Instead, emphasizing the scientific consensus in favor of COVID-19 vaccines and avoiding false balance in communication may help ameliorate misconceptions [Bartoš et al., 2022, Ceccarelli, 2011]. Further, perceived experts and their professional organizations may build trust and disseminate health information more effectively by developing networked communication strategies to rapidly, openly, and factually address false claims that gain traction while clearly explaining areas of uncertainty and directly addressing legitimate safety concerns (as several of the most central perceived experts in the pro-vaccine community did during the study period, see 7.4) [Carpiano et al., 2023]. User-provided expertise cues were sufficient to garner greater engagement on Twitter in vaccine-related discussions, suggesting that individuals could misrepresent their own credentials to more effectively spread anti-vaccine misinformation (Figure 5). Platforms may counter potential deception by establishing a mechanism to verify academic and professional credentials and creating signals within profiles to identify authorities on health-related topics. These measures were implemented on Twitter in the early months of the COVID-19 pandemic, but have since been abandoned. Finally, this work illustrates the importance of self-regulation within expert communities, particularly as medical boards clarify that health professionals who spread vaccine misinformation may face disciplinary consequences [Federation of State Medical Boards Ethics and Professionalism Committee, 2022, SoRelle, 2022].

## Limitations and Extensions

This study relies on proxies for tweet content and user activity that may miss important variation and nuance in stance. Breaking the network into pro- and anti-vaccine communities is common across studies of social media networks [Hagen et al., 2022, Sharma et al., 2021, **?**, Jamison et al., 2020], and stance was generally consistent across popular tweets in either community (Figure 8), but there were several notable exceptions. Positive tweets from users in the anti-vaccine community tended to cite vaccine efficacy as an argument against mandating vaccines, while negative tweets from users in the pro-vaccine community (particularly those by perceived experts) praised vaccine regulators for responding to safety signals. Further, sharing a particular misinformation or academic link may not constitute endorsement, particularly in fact-checking contexts. Although academic sources were shared in the anti-vaccine community, academic sources may be misrepresented by these users [**?**Beers et al., 2022, Kata, 2010] or include articles that have been retracted [Van Schalkwyk, 2019, Abhari et al., 2023]. Future work may examine which academic links were shared within the anti-vaccine community and how these sources were interpreted. More detailed content analysis may reveal important differences between the communities beyond attitudes toward vaccine (e.g. attitudes toward non-pharmaceutical interventions and perceptions of severity of COVID-19 infection), heterogeneity in vaccine opinion within groups, and specific rhetorical strategies utilized by perceived experts in either group [Bradshaw et al., 2020, Maddox, 2022].

An important limitation of our study is that we were unable to assess how many users are exposed to a given tweet. By relying on engagements as an indicator of tweet popularity, we may underestimate the true reach of content. We also could not ascertain how exposure to vaccine-related information in this study influenced health decision-making and behavior, questions that could be directly evaluated in an experimental setting [Jiménez et al., 2018]. Further work may additionally examine the relative contribution of prestige bias to vaccine decision-making compared to other types of biases and how the effects of perceived expertise are moderated by other source characteristics (e.g., trust-worthiness and competence) [Pornpitakpan, 2004] and other forms of bias (e.g., confirmation bias, repetition bias, or success bias) [Acerbi, 2016, Jiménez et al., 2018, Brand et al., 2020, Berl et al., 2021, Nadarevic et al., 2020, Lachapelle et al., 2014, Lin et al., 2016].

This analysis is limited to a single month, based on events in the United States and English keywords, constrained to Twitter, and focused on conversations about COVID-19 vaccination. Initial vaccination capacity, trust in experts, and vaccine uptake vary considerably between countries [Wilson and Wiysonge, 2020] and our own analysis found considerable geographically clustering across the coengagement network (Figure 10). Further work may examine how the role of perceived experts in conversations about vaccination differed between regions and across different time periods (including prior to the COVID-19 pandemic and after the emergence of variants of concern with high breakthrough infection rates). Although we excluded the relatively small set of individuals who added or removed signals of expertise from their profile during the month of the study, extending the study period to expand this set of users could enable further examination of how user behavior and influence changes depending on perceived expertise [Hasan et al., 2022]. Twitter users are not representative of the general population [Wojcik and Hughes, 2019], and patterns in vaccine conversations on social media do not necessarily reflect actual vaccination trends [Cascini et al., 2022, Tavoschi et al., 2020]. The generalizability of these findings should be assessed on different social media platforms [Jones et al., 2012, Cascini et al., 2022, Tangcharoensathien et al., 2020]. The role of experts, particularly those who take counter-consensus positions, is relevant across other scientific topics including climate change, tobacco, and AIDS etiology and treatment [Oreskes and Conway, 2022, Epstein, 1996]. These methods could be applied to compare the role of perceived experts in conversations about different scientific and non-scientific topics (e.g., politics and entertainment) to test the extent to which domain specific credentials are necessary to be perceived as an experts and compare the effects of perceived expertise relevant to different conversations (e.g., whether politicians speaking on political matters receive an influence boost comparable to that of medical professionals and scientists discussing COVID-19 vaccines).

## Conclusion

We found that the set of anti-vaccine perceived experts extends far beyond prominent examples noted by others previously [Smith, 2017, The Virality Project, 2022], suggesting that they should be addressed as a unique and sizable group that blends misinformation with arguments that appear scientific. We also found evidence that perceived experts are more influential than other individuals in the anti-vaccine community, as they disproportionately occupied central network positions where they could reach large audiences and received significantly more engagements (20% more likes and 37% more retweets) on their vaccine-related posts compared to perceived non-experts. Perceived experts are not only some of the most effective voices speaking out against vaccine misinformation; they may be some of its most persuasive sources.

## 5 Methods

### 5.1 Data collection

The Institutional Review Board of Washington University determined that this study (STUDY00017030) was exempt. Our analysis was conducted on a subset of collections of public tweets containing keywords related to vaccines and the COVID-19 pandemic, retrieved and stored in by the University of Washington Center for an Informed Public in real-time as they were posted. Tweets that were later deleted and public tweets from accounts that were suspended or became private are included in the dataset.

We constrained our search period to April of 2021, noting that all individuals sixteen and older were eligible for vaccination by April 19th, marking this time period as an especially critical window for vaccine decision-making [Roy, 2021]. Further, administration of the Johnson & Johnson (Janssen) vaccine was paused in the United States between April 13th and April 23rd while the Centers for Disease Control and Prevention (CDC) and Food and Drug Administration (FDA) investigated a safety signal involving six reported cases of severe blood clots [U.S. Food and Drug Administration, 2021]. Focusing on April of 2021 also allows us to examine how different communities reacted to credible news of a serious but rare vaccine safety signal. During the same time period, fact checkers and researchers responded to several false claims about vaccination, including rumors that vaccinated people were able to “shed” vaccine components that might infect and harm unvaccinated people [The Virality Project, 2021].

To focus our analysis on content related to COVID-19 vaccines, we selected tweets (i.e., posts) within the collections that mentioned keywords related to vaccines in general or the specific manufacturers of the three COVID-19 vaccines initially offered in the United States (Supplemental File 1). Although this protocol focuses on vaccine administration in the United States and English language, tweet selection was not constrained to a specific geographic region. In total, we retrieved 4,276,842 unique tweets including quote tweets (when another post is shared with commentary) and replies (a direct response to another user’s tweet) from April. 5,523,595 unique users participated in the Twitter conversation about COVID-19 vaccines during the study period by either posting original content or retweeting (i.e., sharing) another user’s tweet on the topic. We additionally retrieved retweets, quote tweets, and replies linked to tweets in the April collection that were posted within 28 days of the original tweet (extending the dataset to May 28th) to compare the number of likes and retweets each tweet in the April study period received across a window of the same length (four weeks).

We randomly generated a unique numeric identifier for each user to protect user privacy (particularly for users who are not public figures) while allowing our findings to be reproduced. Individual users are not named in the analyses, reinforcing that we aim to characterize the *group* of perceived experts within the anti-vaccine community instead of focusing on individual, high-profile examples. User profiles and numeric identifiers for accounts used by Twitter were stored separately from tweets, which were instead linked to the anonymous identifiers we assigned. We also stored a dictionary that allows for translation between these two distinct numeric identifiers and utilized the dictionary to connect statistics about tweets to properties of the network and users (e.g., to determine community stance after tagging the stance of a sample of popular tweets, to compare the types of links shared depending on perceived expertise and community, and to collect matching covariates based on Twitter activity and engagements). In tweets we coded for stance, we anonymized mentions of other usernames and removed links to other posts and images on Twitter to prevent user identification. Therefore, researchers were blinded to user identity when assessing tweet stance. We have made tweet and user information relevant to results in this study publicly available without the linking dictionary.

### 5.2 Coengagement network construction

We examined the activity of influential users with shared audiences by generating a coengagement network. First, we constructed a directed graph where each edge connects a user to another user whom they have retweeted at least twice. Next, we used a Docker container developed by Beers et al. [2023] to transform the network into an undirected graph. Edges link accounts that were retweeted at least twice by at least ten of the same users. The resulting graph therefore filters users to those that received some amount of repeated engagement from several accounts rather than those that produced a single viral tweet. We used the Infomap hierarchical clustering algorithm implemented at mapequation.org to detect communities within the coengagement network. Infomap balances the detection of potential substructures (i.e., subcommunities within larger communities) against concisely describing a random walker’s movements through the network to determine the total number of levels to use [Rosvall and Bergstrom, 2008, Holmgren et al., 2022]. The subcommunities detected using Infomap correspond well to communities detected using the Louvain method, an alternative community detection algorithm (Supplemental Figure 10, Supplemental Figure 11, Supplemental Table 4). Coengagement networks were visualized using the open-source software package Gephi with the ForceAtlas2 layout algorithm [Bastian et al., 2009, Jacomy et al., 2014].

### 5.3 Tagging profiles

Based on username, display name, and user description collected each time a user tweeted or was retweeted during the study period, we manually labeled each profile in this group as a perceived expert or not (Supplemental Figure 7). The lead author tagged all users in the coengagement network and two additional authors labeled a subset of 500 users to test robustness of tags based on interrater reliability. We first noted whether a profile indicated that the user primarily tweeted in a language other than English. Because we were unable to assess non-English expertise signals and tweets associated with these users, who may also have reached a substantially different audience compared to English-language accounts, we excluded non-English accounts from our analysis. We also noted accounts that appeared not to represent individuals (e.g., accounts for media groups, non-profit organizations, governmental agencies, and bot accounts). Non-English and non-individual accounts (2,549 accounts in total) remained in the network for visualizations and calculations related to network centrality but were excluded from analyses focused on comparing individuals with and without signals of expertise in their profiles.

For the remaining individual profiles, we noted whether there were signals of expertise in the account username or display name (which are listed with tweets in a user’s feed) or the account description (which is only visible if a user mouses over the author’s tweet or directly visits the account) (Supplemental Figure 7). Signals of expertise included academic prefixes (e.g., Dr. or Professor), suffixes (e.g., MD, MPH, RN, PhD), and professional information (e.g., scientist, retired nurse). We limited our definition of perceived expertise to include training or professional experience in a potentially relevant field but excluded individuals who expressed an interest in a related topic without providing qualifications (e.g., “virology is the coolest”). We included anonymous accounts and ones that may have been parodies (e.g., “Dr. Evil” and “The Mad Scientist”) since we expect that users evaluate profiles based on heuristics and without investigating the veracity of information provided [Sundar, 2007]. We assumed that medical and wellness professionals, including practitioners of alternative medicine, may broadly be perceived as experts regardless of specialty [Jiménez and Mesoudi, 2019]. To focus on biomedical expertise, we did not code users from other fields as perceived experts (e.g., science journalists, disability rights advocates, and governmental officials without biomedical backgrounds), acknowledging that these sources may provide trusted and knowledgeable perspectives relevant to health decision-making.

Users that indicated expertise in any part of their profile (names, description, or both) at any point in time were tagged as perceived experts based on experimental evidence that users with biomedical expertise signals in their profiles are perceived to have greater expertise on COVID-19 vaccines [Jalbert et al.]. Users that changed their profiles over the course of the study in a manner that changed whether they might be perceived as an expert (24 accounts) were excluded from the following analyses.

### 5.4 Determining community stance

To understand the stance of individuals in the two largest communities, we analyzed tweets that received retweets during April 2021. Of the 251,040 tweets satisfying those criteria, we tagged a subset posted by the ten perceived experts and perceived non-experts in both communities with the greatest degree centrality. We collected the ten most retweeted tweets by the forty selected users. A few of the users had fewer than ten tweets that were retweeted during the study period, so we instead retrieved all of their tweets that received retweets in April. In total, we reviewed 392 tweets.

For each tweet, three coders assessed stance toward COVID-19 vaccines as positive, negative, or neutral. Stance was evaluated as positive if the tweet supported vaccination, including by providing evidence that COVID-19 vaccines are safe and effective. Tweets expressing the opposite view were coded as negative. Stance was often implicit. For example, calls for deployment of vaccines to “hotspots” with elevated disease burden suggest that the author believes the vaccine can prevent disease, even if this rationale is not stated directly. Unless the author refuted or negatively reacted to a quote they provided, a tweet they quote tweeted, or an article they linked, the post was assumed to take the same stance as referenced sources. Tweets containing multiple contrasting viewpoints or providing information without a clearly implied position (e.g., sharing statistics about the pace of vaccination without any additional commentary) were coded as neutral. The neutral tag was also applied to tweets if the coder was uncertain about the argument being made or context was missing (e.g., criticism of an article that could not be retrieved). We noted many tweets during this time period that opposed vaccine mandates and coded these tweets as neutral unless the author justified their position using a specific argument regarding vaccine safety or efficacy. Tweets comparing multiple types of vaccines with negative stances toward some and positive stances toward others were tagged neutral. Tweets that took a uniform stance toward different types of COVID-19 vaccines while explaining their differences were coded based on the corresponding stance. Based on this analysis, we determined that negative stances toward COVID-19 vaccines were prevalent in tweets from one community, which we refer to as the anti-vaccine community in following analyses (Supplemental Figure 8). The other community, in which tweets mainly expressed positive stances, was the pro-vaccine community that served as a basis of comparison.

### 5.5 Academic and low-quality link-sharing

We assessed the types of sources users referenced in their tweets by comparing links shared to databases of: (1) low quality sources and (2) academic research journals and pre-print servers. We expanded shortened URLs using the RCurl package in R [Temple Lang, 2023]. In some cases, this process timed out or links connected to other content shared on Twitter, in which case links were excluded from the following analysis. Across community and perceived expertise, we compared the average number of tweets per user, proportion of tweets with links, and proportion of links that were checked, and number of links that were checked to ensure there was minimal risk of differences in link-sharing behavior leading to bias in the following analyses (Supplemental Figure 9). We checked the remaining 52,073 links, first determining whether the domain name was rated “low” or “very-low” quality by Media Bias/Fact Check according to the Iffy Index of Unreliable Sources [Golding, 2023, Van Zandt], a common proxy for misinformation sharing [Sharma et al., 2021, DeVerna et al., 2022]. To assess sharing of academic research, we also checked links to research publications [Priem et al., 2022] and pre-print servers used in biomedical and medical sciences [Kirkham et al., 2020]. For perceived experts and perceived non-experts in each of the two large communities, we calculated: (1) the proportion of assessed links from low quality or academic research sources and (2) the proportion of total users that shared at least one link from a low quality or academic research source in their original posts during April 2021. For each metric, we calculated the binomial proportion 95% confidence interval as 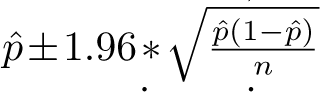 where *p*^ is the observed proportion and *n* is the total links or users in a given category. To compare proportions for different categories of users, we conducted two proportion one-tailed Z-tests with Yates’ continuity correction to account for the small number of links from low quality sources shared by the pro-vaccine community.

### 5.6 Network centrality and bridging metrics

We calculated degree, betweenness, and eigenvector centrality to describe network position using the igraph package in R [Csardi and Nepusz, 2006] (see Supplemental Table 1 for a description of centrality metrics and their interpretation). Specifically, we calculated eigenvector centrality using the PageRank algorithm. We additionally detected nodes with audiences spanning the anti- and pro-vaccine communities using a community bridging index calculated as the minimum number of edges that a node has linking it to either of the two communities.

We tested whether perceived experts were overrepresented within the group of highly ranked users by each metric by calculating the proportion of perceived experts in the top *n* users ranked by a given metric (where *n* varies between 500 and 50 for all metrics and ten for bridging score). Several perceived experts and perceived non-experts had the same bridging scores, leading to ties for ranking in the top 500 and 50 bridges. In these cases, we randomly drew 1,000 samples from the tied users without replacement to complete the set of top bridges and calculated the mean proportion of perceived experts across all samples. To calculate the 95% confidence interval in these cases, we added the margin of error 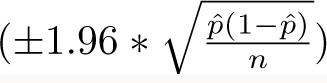 to the .025th and 97.5th percentile proportion value across the 1,000 samples. For the remaining metrics and subsets, we calculated the binomial proportion 95% confidence interval as in the previous section except when examining the ten top bridges, in which case we calculated the Wilson score interval to account for the small sample size.

To test whether perceived experts were significantly overrepresented in each sample of highly ranked individuals, we conducted two proportion Z-tests with the alternative hypothesis that perceived experts were a significantly greater proportion of the top *n* individuals than individuals in the complement (i.e., individuals who were not among the top *n* individuals ranked by a given metric). When considering the top ten bridges, we instead used a Fischer’s exact t-test to account for the small sample size. We reported p-values for significance tests.

### 5.7 Matching to assess relative influence of perceived experts

This portion of the analysis was pre-registered on OSF Registries prior to hypothesis testing (https://osf.io/6u3rn). We assessed how perceived expertise affects influence using propensity score matching, a technique to select a sample of individual perceived non-experts with a similar covariate distribution to perceived exerts in the dataset [Rosenbaum and Rubin, 1983]. We can then compare the average influence between perceived experts and perceived non-experts while adjusting for confounding from covariates that may predict being a perceived expert without being a direct consequence of perceived expertise. Propensity score matching has been applied previously to compare the effectiveness of different users in the context of online advertising [Yan et al., 2018, Kuang et al., 2018] and to understand whether following prominent public health institutions on Twitter impacts vaccine sentiment [Rehman et al., 2016]. Propensity score matching was performed using the MatchIt package in R [Ho et al., 2011].

We matched on the following covariates: natural logged follower count at the beginning of April 2021, total on-topic posts during the study period (i.e., posts included in our data set because they contained a COVID-19 vaccine keyword), account creation date, whether the account is verified, percent of on-topic posts containing links, percent of on-topic tweets that were original posts versus retweets, posting time of day, and uniformity of posting date across the study period (Supplemental Table 2). For the first two analyses focused on relative influence within the anti-vaccine and pro-vaccine communities, we also matched on subcommunity assignment based on the Infomap community detection algorithm Figure 10. We used logistic regression to compute propensity score as the predicted probability that each user is a perceived expert given their covariates. Based on propensity scores, each perceived expert was matched with replacement to their three nearest neighbor perceived non-experts. To test the first two hypotheses about influence within the anti-vaccine and pro-vaccine communities respectively, we included only users in either community. To test the third hypothesis comparing the influence of perceived experts in the anti- versus pro-vaccine community, we calculated the interaction of perceived expertise and community. Sample sizes for all analyses are provided in Supplemental Table 5. The outcome variables for influence are: eigenvector centrality, degree centrality, betweenness centrality, natural logged median likes, natural logged median retweets, h-index for likes, and h-index for retweets (Supplemental Table 3). h-index is the maximum number *h* such that the user wrote at least *h* original tweets (including replies and quote tweets) that received at least *h* likes or retweets. We calculated engagements (likes and retweets) for each tweet as the maximum number observed for that tweet in our collection within twenty-eight days of when it was posted. Importantly, we only retrieved like counts for a given tweet when another user engaged with the tweet through a quote tweet or retweet.

We then estimated the effect of perceived expertise on influence by calculat-ing the average treatment effect on the treated (ATT), which corresponds to the difference in expected influence ((*Y* (*T*)) with and without perceived expertise given that a user is a perceived expert (*T* = 1) with covariates drawn from the distributions for the matched users (X):

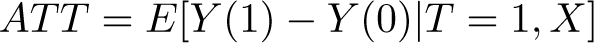

ATT was estimated using the marginal effects package [Arel-Bundock, 2023]. We tested the robustness of our results to different matching parameters and covariates, as displayed in Supplemental Figure 22, Supplemental Figure 23, and Supplemental Figure 24.

## 6 Author information

**Funding:** MJH was funded by the Achievement Rewards for College Scientists Scholarship, the National Institutes of Health (R35GM133439), the University of Washington’s Center for an Informed Public, and the John S. and James L. Knight Foundation. RM was funded by the Bryce and Bonnie Nelson Fellowship. EAM was funded by the National Science Foundation (DEB-2011147, with the Fogarty International Center), the National Institutes of Health (R35GM133439, R01AI168097, and R01AI102918), the Stanford King Center on Global Development, Woods Institute for the Environment, Center for Innovation in Global Health, and the Terman Award. JDW was funded by the Knight Foundation and the National Science Foundation grants 2120496 and 2230616.

**Conflicts of interest**: All authors declare that there are no competing interests.

**Data and code availability:** Code and data used to conduct main analyses and reproduce figures are available on Github at: https://github.com/mjharris95/perceived-experts

As explained in the repository, users were anonymized throughout the analysis and in the dataset. Additional information may be provided by the corresponding author on reasonable request.

## Data Availability

Code and data used to conduct main analyses and reproduce figures are available on Github at: https://github.com/mjharris95/perceived-experts. As explained in the repository, users were anonymized throughout the analysis and in the dataset. Additionally information may be provided by the corresponding author on reasonable request.

## Acknowledgments

We thank Lia Bozarth and Alex Lodengraad for their work constructing and maintaining the Twitter collections used in these analyses, and we thank Lia Bozarth for querying the collection on our behalf. We thank Andrew Beers and Joey Schafer for their assistance with coengagement network methods. We thank Stephen Prochaska for his input on methods to code profiles and tweets and assess interrater reliability. We thank Maddy Jalbert, Luke Williams, Noah Louis-Ferdinand, Alexander Sherman, Caroline Glidden, Desire Nalukwago, Eloise Skinner, Emma Southworth, Isabel Delwel, Jasmine Childress, Kelsey Lyberger, Kimberly Cardenas, Lisa Couper, Marissa Childs, and Mauricio Cruz Loya for their helpful feedback on the manuscript. We additionally thank İhsan Kahveci, members of the James Holland Jones’ lab, and members of the University of Washington’s Center for an Informed Public for their feedback and insights on this project.

## 7 Supplemental Materials

### 7.1 Coengagement network visualization without annotations

**Figure 6:**
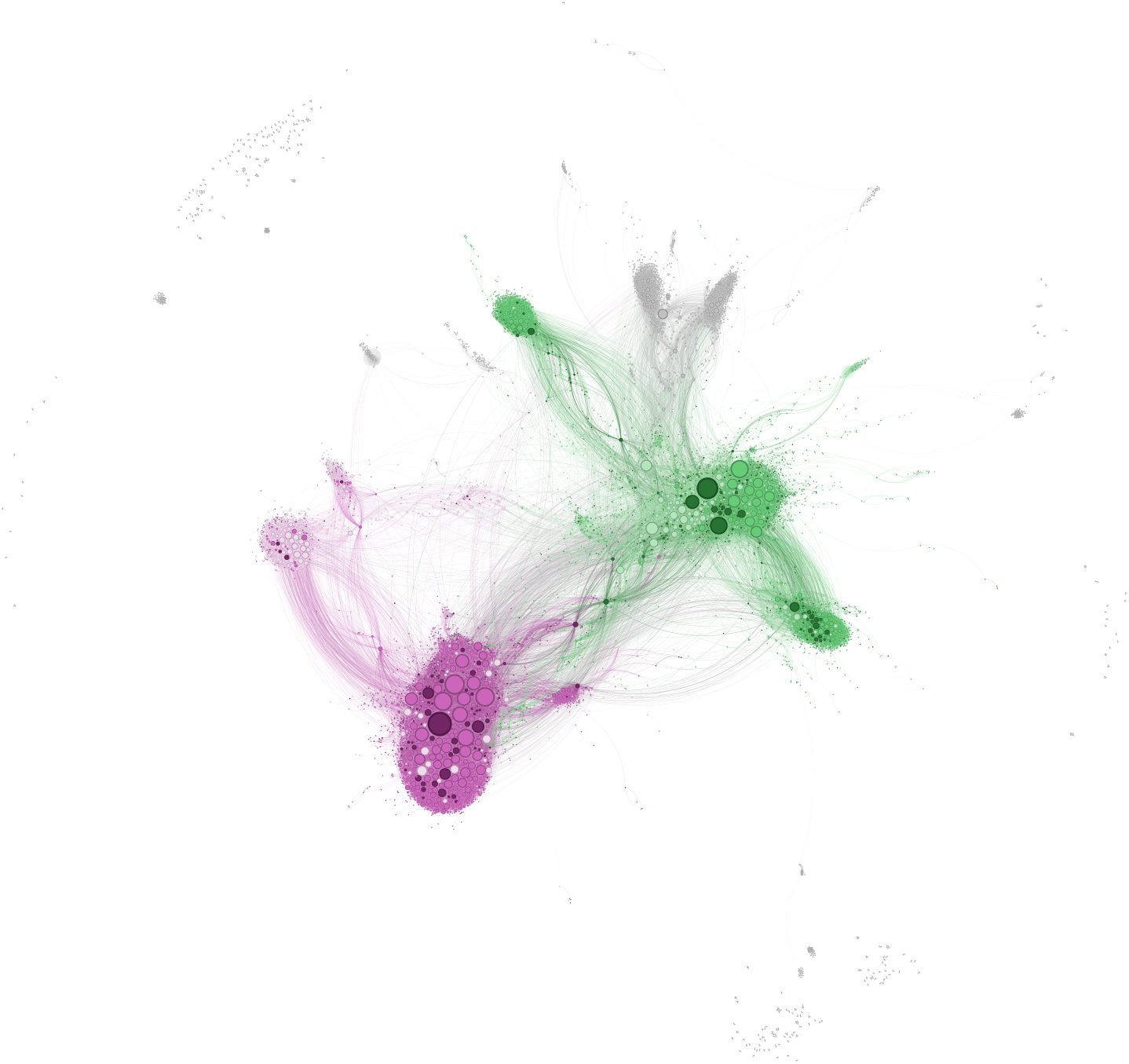
The coengagement network of users tweeting about COVID-19 vaccines is divided into two large communities. Users are represented as circles and scaled by degree centrality. Edges connect users that were retweeted at least twice by at least ten of the same users. Nodes in the two largest communities detected using the Infomap algorithm are colored in pink (anti-vaccine) and green (pro-vaccine). Shades indicate account type: non-individual and non-English accounts excluded from analyses (light); individual perceived non-expert (medium); and perceived experts (dark). Nodes outside of the two largest communities are gray. Each edge is colored based on the color of one of the two nodes it connects, randomly selected. Note that this figure is identical to Figure 1 without annotations and in higher resolution.

#### 7.2 Keywords

Supplemental File 1: Keywords used to search collection for relevant tweets. The asterisk () indicates a wildcard meaning a set of alphabetical characters of any length.

- vacc*
- vaxx
- jab
- shot
- immuniz*
- dose
- mrna
- pfizer,
- j&j
- jnj
- j & j
- j n j
- j and j
- johnson and johnson
- johnson & johnson
- janssen
- moderna

#### 7.3 Variable definitions

This section provides explanations of variables used in our analyses, including centrality metrics (Supplemental Table 1) and matching covariates (Supplemental Table 2) and influence metrics used in propensity score matching (Supplemental Table 2).

**Table 1:** Description of different centrality metrics and their interpretation in the context of coengagement networks.

| Metric name | Description | Coengagement interpretation |
| --- | --- | --- |
| Degree centrality | Number of edges connecting a given node to other nodes | Selects users who share an audience with many other users; proxy for audience size |
| Betweenness centrality | Number of times a given node appears along the shortest path between two other nodes in the network | Selects users who share audience with users that otherwise do not have much overlap in their audiences |
| Eigenvector centrality | Recursively assigns nodes a value based on whether they are connect to other nodes with high eigenvector centrality | Selects users who share audiences with other users whose audiences overlap with those of many others in the coengagement network |
| Community bridging | Takes the minimum of the number of edges that a node has connecting it to nodes in any focal communities | Selects users who share an audience with relatively many users in both the anti- and pro-vaccine communities |

**Table 2:** Description of covariates used for matching.

| Matching covariates | Description |
| --- | --- |
| Creation date | Continuous variable. Date and time that account was created. |
| Follower count (log) | Continuous variable. Natural log of the earliest follower count retrieved for a user within the dataset. |
| Account verified? | Binary variable. Whether account was verified at any point in April of 2021. Note: twelve users in the dataset, eight of whom were in the two largest communities, became verified during the study period.” |
| On-topic post count (log) | Continuous variable. Natural log of the total number of posts (including quote tweets and replies) and retweets by a user in April containing COVID-19 vaccine keywords. |
| Percent retweets | Continuous variable. Percent of user’s on-topic posts in April that were retweets. |
| Percent with links | Continuous variable. Percent of user’s on-topic posts in April that contained links. |
| Morning poster? | Binary variable. Whether at least one third of a user’s on-topic posts were between 6 AM and noon Eastern Time. |
| Afternoon poster? | Binary variable. Whether at least one third of a user’s on-topic posts were between noon and 6 PM Eastern Time. |
| Evening poster? | Binary variable. Whether at least one third of a user’s on-topic posts were between 6 PM and midnight Eastern Time. |
| Night poster? | Binary variable. Whether at least one third of a user’s on-topic posts were between midnight and 6 AM Eastern Time. |
| Uniform post dates? | Binary variable. Whether the dates of a user’s on-topic post are uniformly spaced across the study period. Calculated by conducting a chi-squared test comparing the distribution of post dates to a uniform distribution and testing whether the resulting p-value is less than 0.10. |
| Subcommunity 1-2 | Binary variable. Whether the user is in subcommunity 1-2 (the largest anti-vaccine subcommunity). Only used to test the first hypothesis (influence within anti-vaccine community alone). |
| Subcommunity 1-1 | Binary variable. Whether the user is in subcommunity 1-1 (the second-largest anti-vaccine subcommunity). Only used to test the first hypothesis (influence within anti-vaccine community alone). |
| Subcommunity 2-1 | Binary variable. Whether the user is in subcommunity 2-1 (the largest pro-vaccine subcommunity). Only used to test the second hypothesis (influence within pro-vaccine community alone). |
| Subcommunity 2-2 | Binary variable. Whether the user is in subcommunity 2-2 (the pro-vaccine subcommunity focused on vaccination in Canada). Only used to test the second hypothesis (influence within pro-vaccine community alone). |

**Table 3:** Description of influence metrics that were outcomes for matching.

| **Outcome variable** | **Description** |
| --- | --- |
| Median likes (log) | Continuous variable. Natural log of median number of likes received on all original (non-retweet) on-topic posts. |
| h-index likes (log) | Continuous variable. Natural log of h-index for likes received across all original (non-retweet) on-topic posts. |
| Median retweets (log) | Continuous variable. Natural log of median number of retweets received on all original (non-retweet) on-topic posts. |
| h-index retweets (log) | Continuous variable. Natural log of h-index for retweets received across all original (non-retweet) on-topic posts. |
| Degree centrality | Continuous variable. Degree centrality within coengagement network. |
| Eigenvector centrality | Continuous variable. Eigenvector centrality within coengagement network. |
| Betweenness centrality | Continuous variable. Betweenness centrality within coengagement network. |

#### 7.4 Tweet and user characteristics

This section contains additional information about expertise signals provided by users (Supplemental Figure 7), the vaccine stances of popular tweets in the the anti- and pro-vaccine communities (Supplemental Figure 8), and link-sharing behavior for users depending on perceived expertise and community (Supplemental Figure 9).

**Figure 7:**
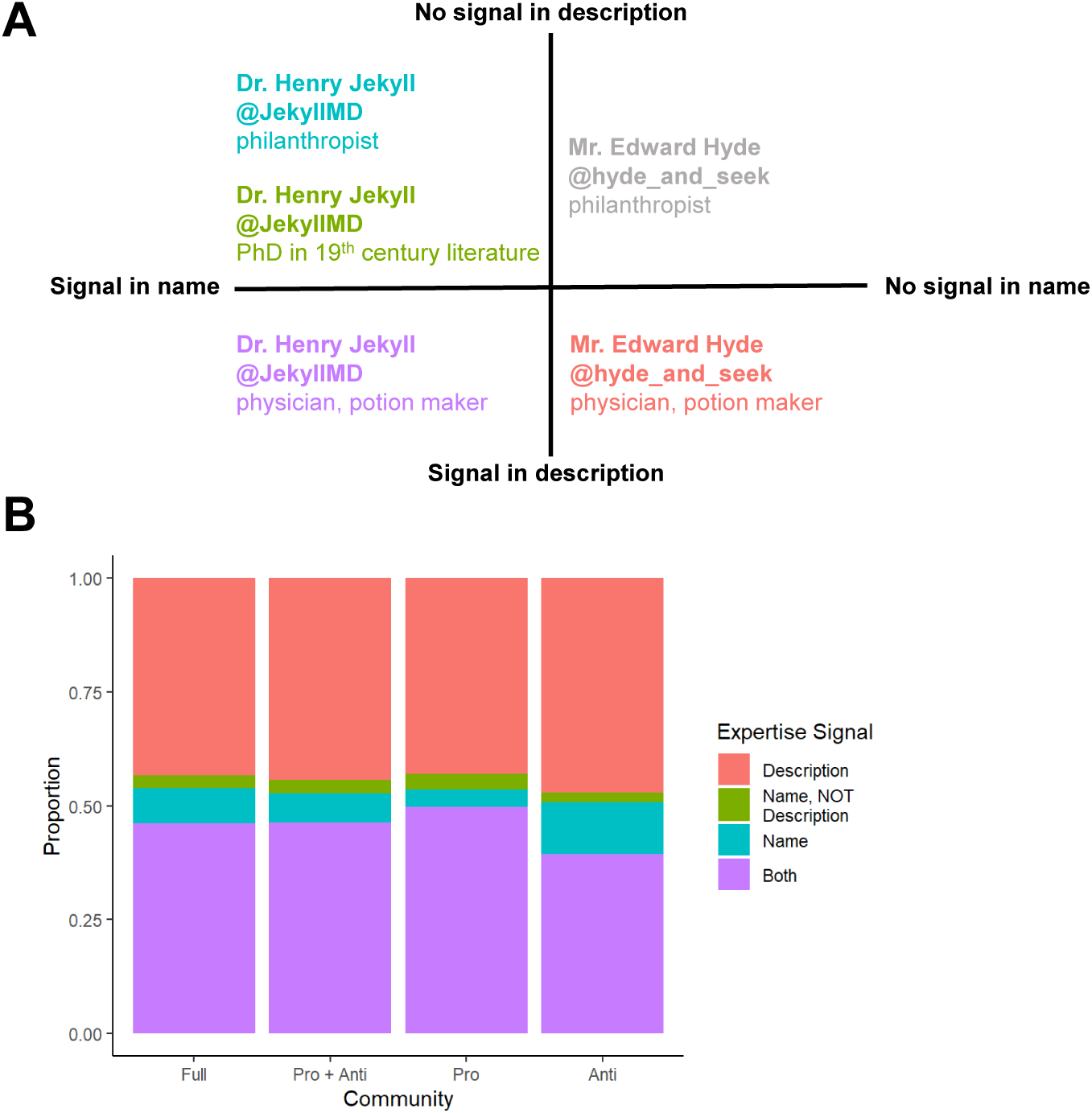
Signals of expertise may appear in different parts of the profile. Panel A (top) gives examples of different ways that expertise signals may be presented. Starting in the bottom right and moving counterclockwise: signal in description only (red), signal in name and description (purple), signal in name but description clarifies user is not an expert (green), or signal in name only no information about perceived expertise in description (blue). The gray profile has no signals of expertise and is therefore not a perceived expert. Panel B (bottom) is the proportion of perceived experts that provide expertise signals in each way across different group (x-axis): the full coengagement network, the pro and anti-vaccine communities combined, the pro-vaccine community alone, and the anti-vaccine community alone.

**Figure 8:**
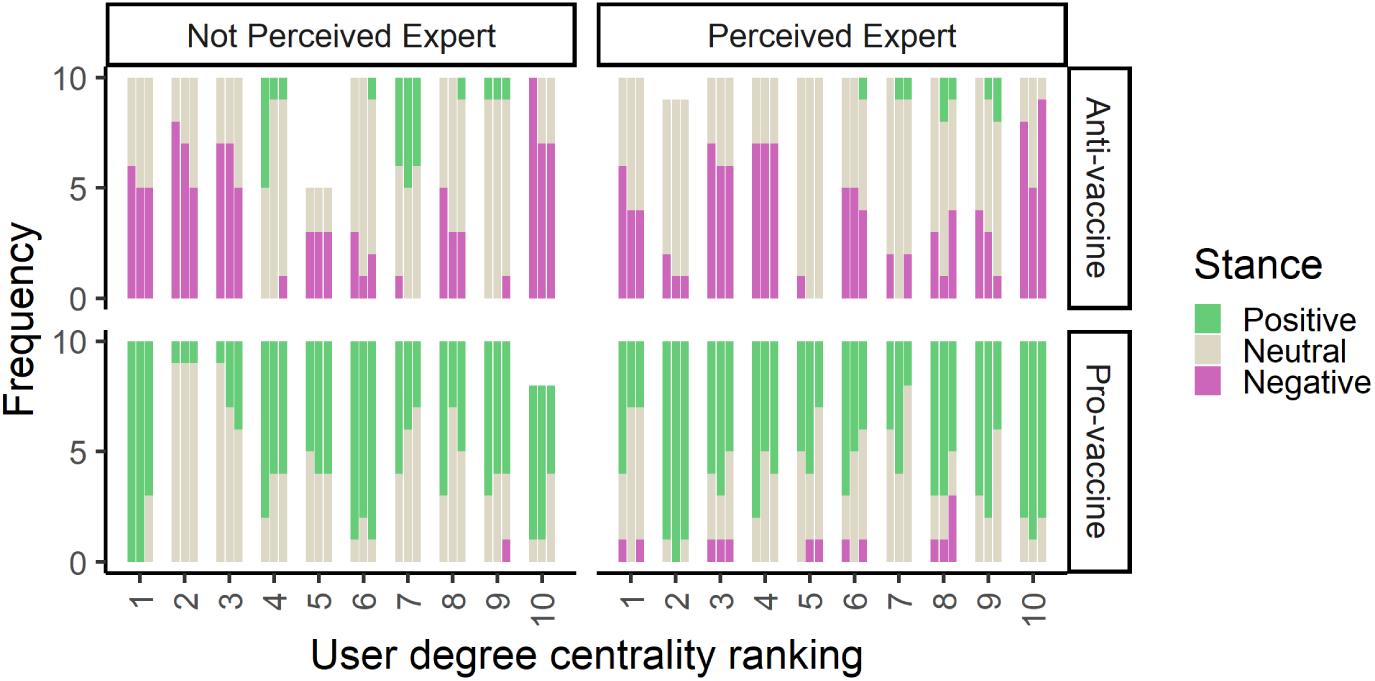
User stance is largely consistent within the two largest communities. Plots display tweet stance across the ten perceived non-experts (left column) and ten perceived experts with the greatest degree centrality (right column) in the anti- and pro-vaccine communities (top and bottom row respectively). Users are ranked and arranged by degree centrality (x-axis). Three coders assessed the stance of a sample of highly retweeted tweets from each user, and results from all coders are displayed side-by-side for each user. Bars are shaded by the number of tweets assessed as positive (green), neutral (gray), or negative (pink).

**Figure 9:**
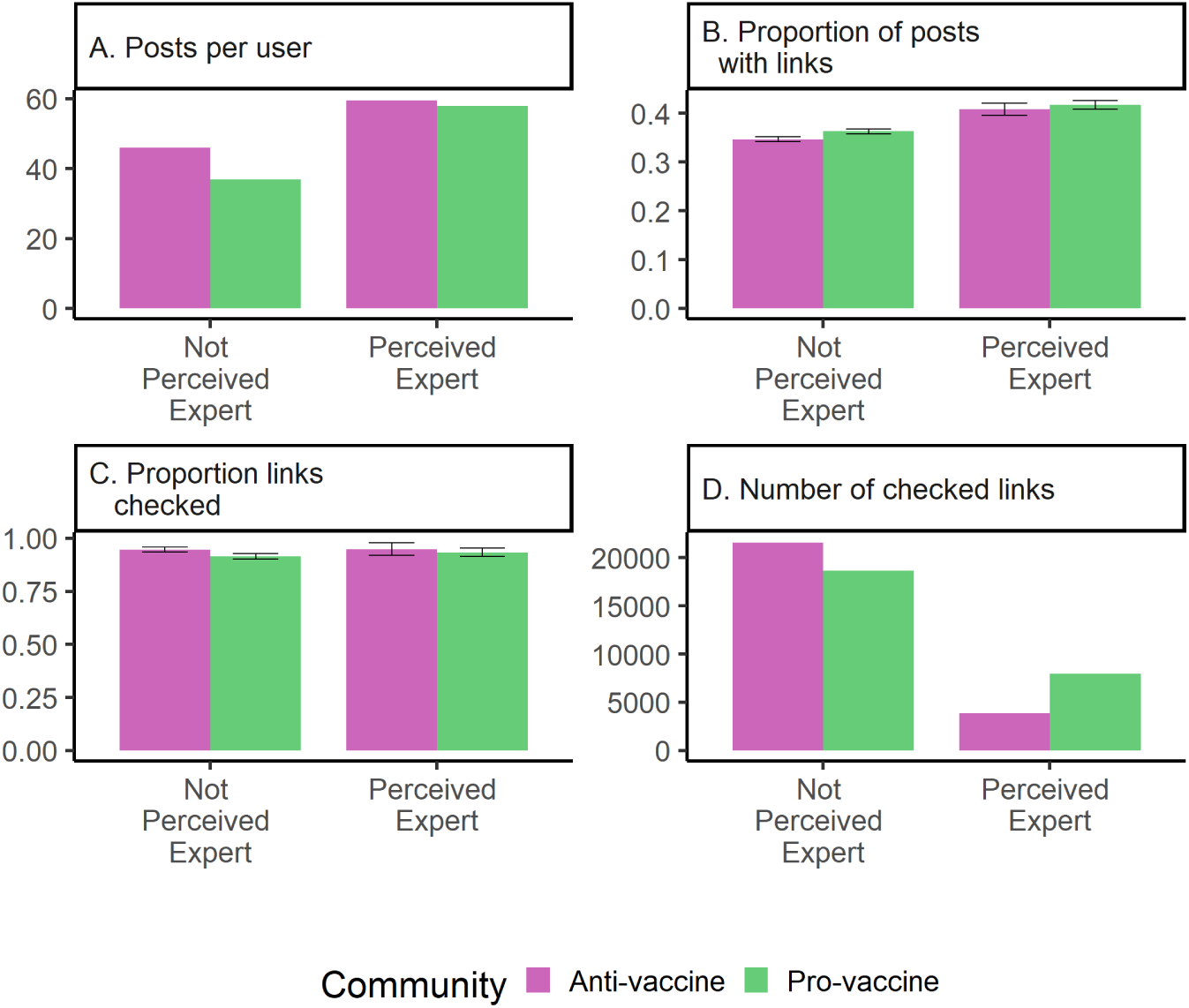
Link-sharing activity across communities and perceived expertise. Each panel compares a different metric of link-sharing by perceived experts and perceived non-experts in the anti- (pink) and pro- (green) vaccine communities. The metrics are: (A) average number of posts per user (B) proportion of all tweets with links (C) proportion of links that were checked (did not link back to Twitter or timed out during link expansion) (D) the total number of checked links in a given category. Error bars give 95% binomial proportion confidence intervals for proportions in panel B and C.

#### 7.5 Subcommunity descriptions and visualization

Infomap is a hierarchical community detection algorithm, allowing the detection of subcommunities [Holmgren et al., 2022]. In this section, we visualize sub-communities detected using Infomap (Supplemental Figure 10) and note that these subcommunities closely resemble communities detected using the Louvain method, suggesting that our results are robust to choice of community detection algorithm. We also provide a table of subcommunity properties (Supplemental Table 4).

We reviewed a sample of popular tweets from each subcommunity to label subcommunities based on topic. Within the pro-vaccine community, the four most popular subcommunities included: the largest (main) subcommunity that generally discussed vaccine safety and efficacy and focused on vaccination in the United States, a subcommunity of predominantly non-individual accounts that posted news stories with headlines, a subcommunity focused on vaccination in Canada, and a subcommunity focused on vaccination in Australia. Tweets in the latter two subcommunities often criticized the pace of vaccination as too slow, blaming specific politicians. The four most popular anti-vaccine subcommunities included two large (main) subcommunities that we labeled as main anti (A) and main anti (B). The distinction between the two subcommunities may be related to main anti (A)’s greater connection to followback clusters, or densely connected groups of accounts that frequently retweet each other and a small set of accounts outside of their network [Beers et al., 2023], although we did not test this hypothesis directly. Again, these subcommunities generally discussed vaccine safety and efficacy, with some emphasis on vaccination in the United States. The two next largest subcommunities in the anti-vaccine community consisted largely of non-English accounts with profiles in French or Italian. The third largest community that was excluded from the main analysis tweeted about vaccination in India ([Hagen et al., 2022, Boucher et al., 2021]), which broke into two subcommunities: India (A) and India (B). It was not immediately clear whether there were any differences in the content posted by users in either subcommunity.

The communities detected using the Louvain method closely resembled these subcommunities, although the media subcommunity in the pro-vaccine community was combined with the main pro-vaccine subcommunity. Across subcommunities, all users were fully contained within their equivalent Louvain subcommunity except in the case of anti (A), as some users in the boundary between anti (A) and anti (B) were classified into the latter subcommunity. There were 2004 users in the largest Louvain communities that were not in any of the largest Infomap subcommunities, meaning that the Louvain communities were generally more expansive.

**Figure 10:**
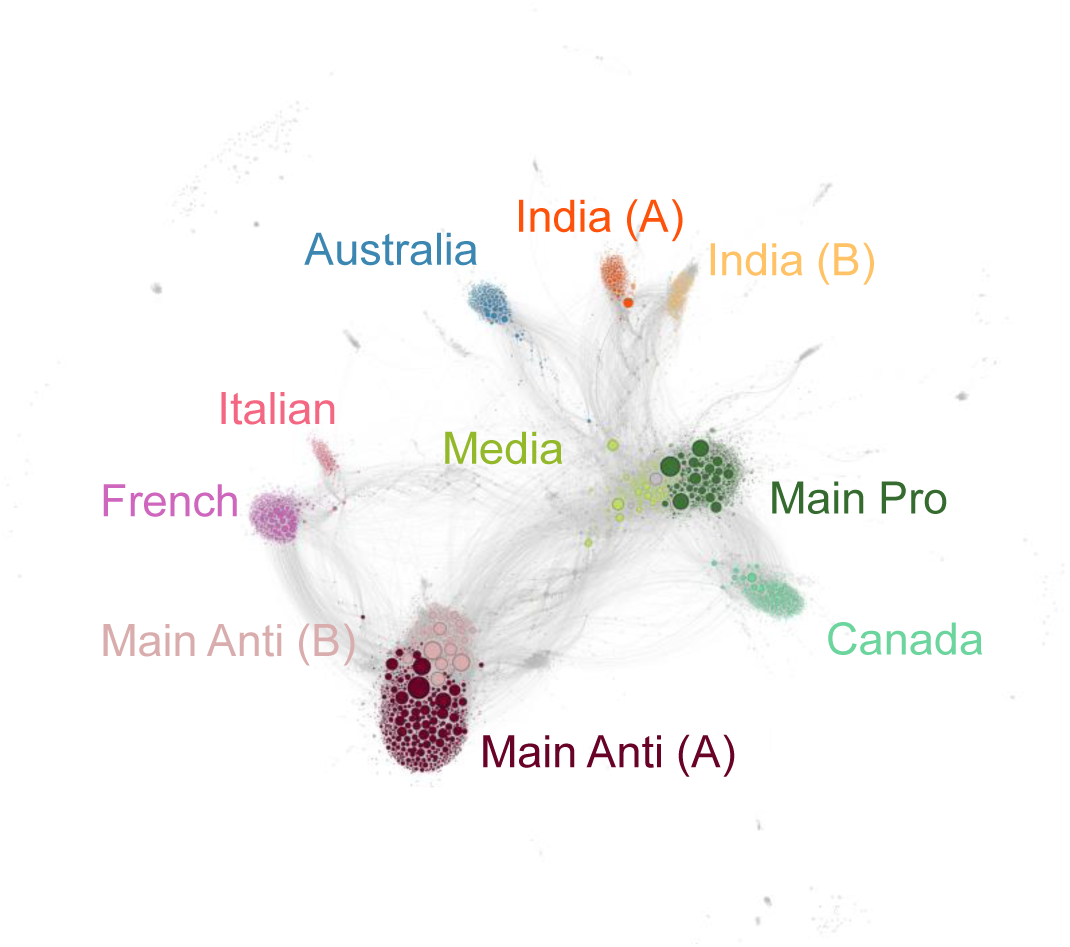
The main subcommunities of the coengagement network of users tweeting about COVID-19 vaccines. Users are represented as circles and scaled by degree centrality. Edges connect users that were retweeted at least twice by at least ten of the same users. All edges are gray. Nodes in the ten largest subcommunities detected using the Infomap algorithm are colored in different shades of pink and red if they are in the anti-vaccine community (top to bottom: Italian, French, Main Anti (B), Main Anti (A)), green and blue if they are in the pro-vaccine community (top to bottom: Australia, Media, Main Pro, and Canada), and shades of orange if they are in the community focused on vaccination in India (top to bottom: India (A), India (B)). Nodes outside of the ten largest subcommunities are gray.

**Figure 11:**
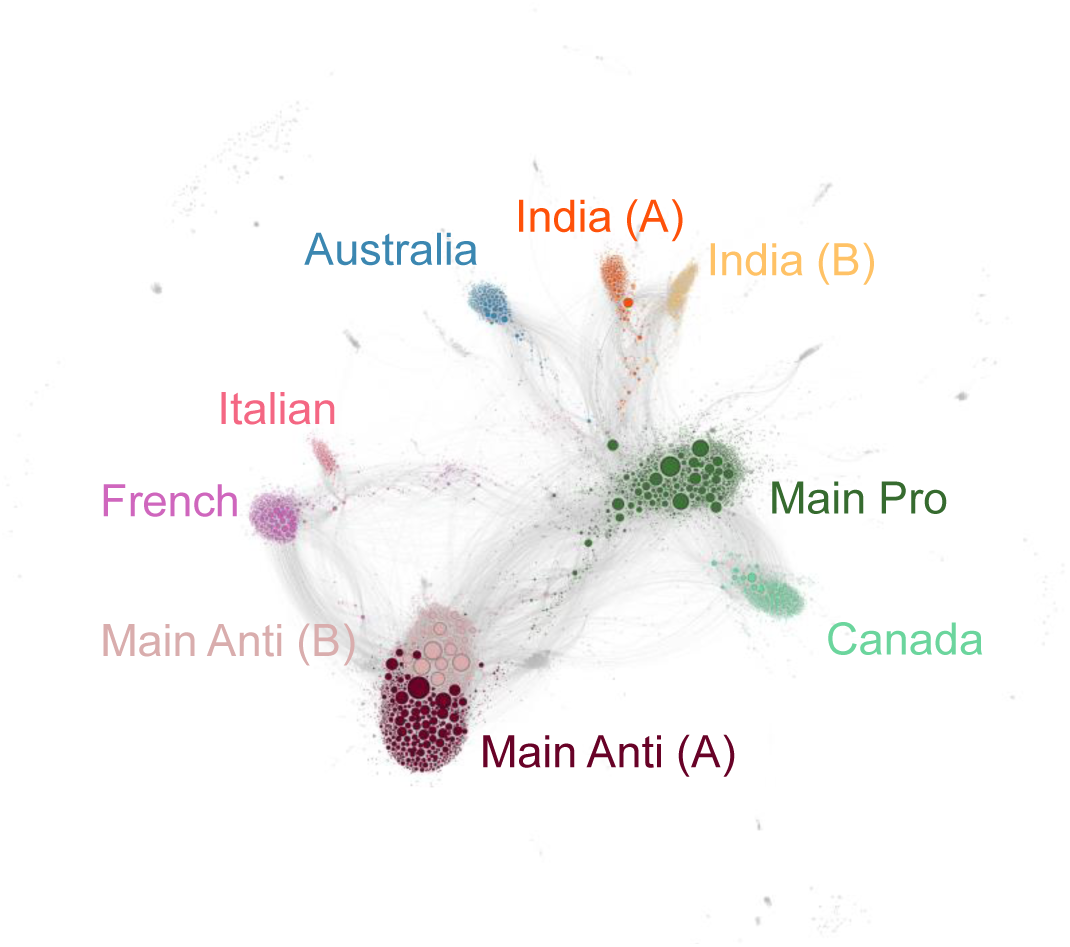
The main communities of the coengagement network of users tweeting about COVID-19 vaccines detected using the Louvain algorithm roughly correspond to subcommunities detected using the Infomap algorithm (compare to Supplemental Figure 10). Users are represented as circles and scaled by degree centrality. Edges connect users that were retweeted at least twice by at least ten of the same users. All edges are gray. Nodes in the nine largest communities detected using the Louvain algorithm are colored in different shades of pink if they are in the anti-vaccine community (top to bottom: Italian, French, Main Anti (B), Main Anti (A)), green if they are in the pro-vaccine community (top to bottom: Australia, Main Pro, and Canada), and shades of orange if they are in the community focused on vaccination in India (top to bottom: India (A), India (B)). Nodes outside of the nine largest subcommunities are gray.

**Table 4:** Properties of subcommunities detected using the Infomap community detection algorithm (displayed in Supplemental Figure 10). From left to right, the columns are as follows: the code for the subcommunity (where the first digit corresponds to the community code and the second digit corresponds to the subcommunity code); a subcommunity name determined by reviewing a sample of popular tweets; the name of the equivalent subcommunity detected using the Louvain community detection algorithm; the name of the community to which the subcommunity belongs; the total number of accounts in the subcommunity and, in parentheses, the proportion of total users in the community that are part of a given subcommunity; the number of users meeting the criteria for inclusion (i.e., English profile, individual, unchanged perceived expertise) and, in parentheses, the proportion of users in the subcommunity meeting the criteria for inclusion in analyses; the number of perceived experts in the subcommunity and, in parentheses, the proportion of individual users in the subcommunity who are perceived experts; and the count of users in a given Infomap subcommunity who are also in the corresponding Louvain community and, in parentheses, the proportion of users in the Infomap subcommunity who are in the corresponding Louvain community.

| Subcom-<br>munity<br>Code | Name | Louvain<br>Equiva-<br>lent | Community | Total<br>users | Included<br>users | Perceived<br>Experts | Count in<br>Louvain |
| --- | --- | --- | --- | --- | --- | --- | --- |
| 1 1 | Anti (A) | Anti (A) | Anti | 744 (0.28) | 684 (0.92) | 76 (0.11) | 662 (0.89) |
| 2 1 | Main Pro | Main Pro | Pro | 737 (0.21) | 579 (0.79) | 84 (0.15) | 737 (1) |
| 1 2 | Anti (B) | Anti (B) | Anti | 538 (0.2) | 491 (0.91) | 40 (0.08) | 538 (1) |
| 1 3 | French | French | Anti | 425 (0.16) | 90 (0.21) | 7 (0.08) | 425 (1) |
| 2 2 | Canada | Canada | Pro | 385 (0.11) | 283 (0.74) | 56 (0.2) | 385 (1) |
| 2 4 | Australia | Australia | Pro | 316 (0.09) | 289 (0.91) | 42 (0.15) | 316 (1) |
| 2 3 | Media | Main Pro | Pro | 258 (0.07) | 49 (0.19) | 8 (0.16) | 258 (1) |
| 3 2 | India (A) | India (A) | India | 239 (0.35) | 194 (0.81) | 17 (0.09) | 239 (1) |
| 3 1 | India (B) | India (B) | India | 186 (0.27) | 162 (0.87) | 17 (0.1) | 186 (1) |
| 1 5 | Italian | Italian | Anti | 183 (0.07) | 50 (0.27) | 2 (0.04) | 183 (1) |

#### 7.6 Network visualization with nodes scaled by different metrics

This section contains visualizations of the coengagement network where nodes are scaled and labeled according to each network metric used in the analyses: community bridging (Supplemental Figure 12), degree centrality (Supplemental Figure 13), betweenness centrality (Supplemental Figure 14), and eigenvector centrality (Supplemental Figure 15). These figures allow comparisons of how different nodes are ranked across each metric.

**Figure 12:**
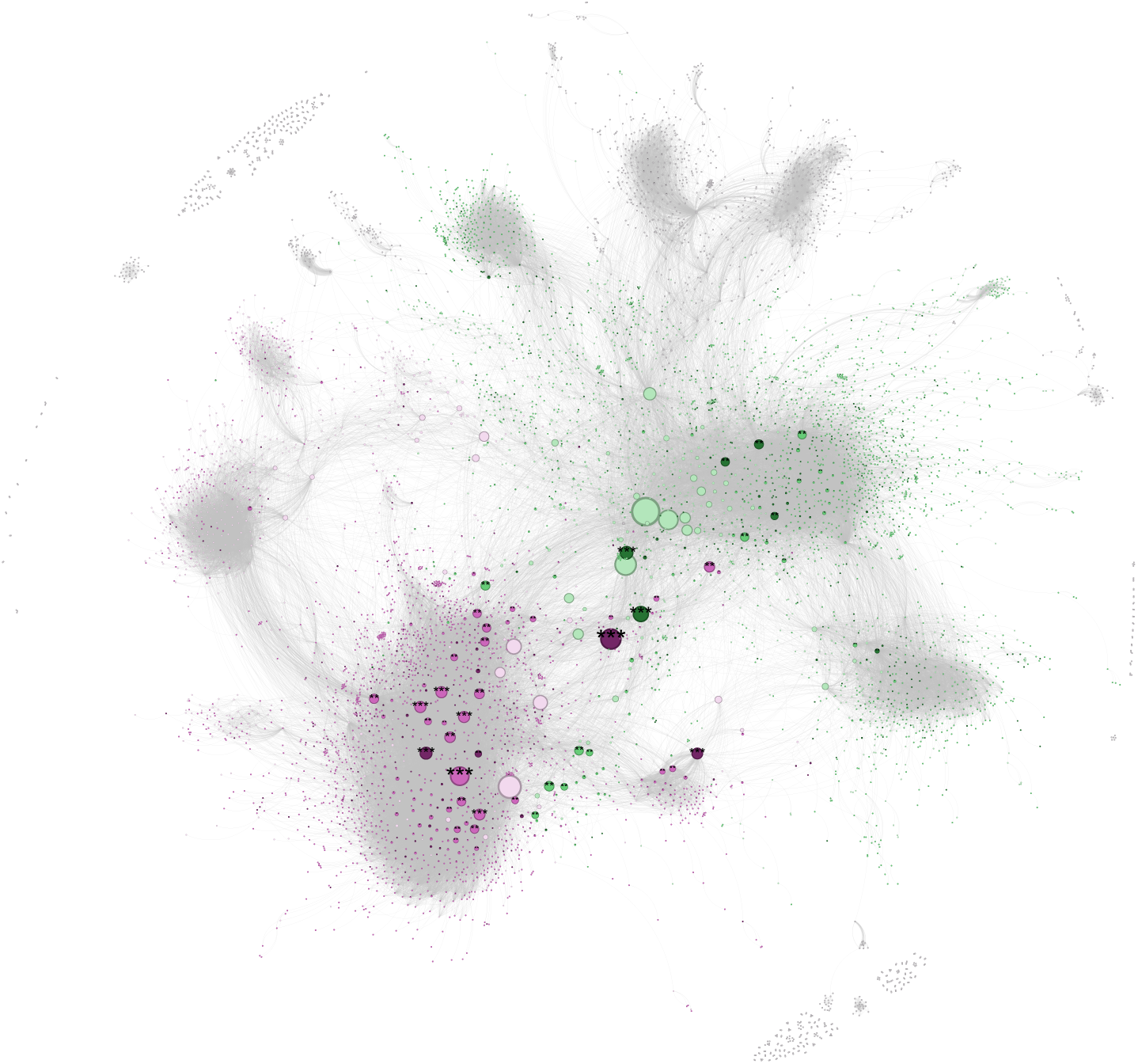
Node size corresponds to *community bridging* score. As in Figure 1, users are represented as circles. Edges connect users that were retweeted at least twice by at least ten of the same users. All edges are gray. Nodes in the two largest communities detected using the Infomap algorithm are colored in pink (anti-vaccine) and green (pro-vaccine). Shades indicate account type: non-individual and non-English accounts excluded from analyses (light); individual perceived non-expert (medium); and perceived experts (dark). Nodes outside of the two largest communities are gray. Users that rank in the top 500, top 50, and top ten users by community bridging across the whole coengagement network are labeled with one, two, and three stars respectively.

**Figure 13:**
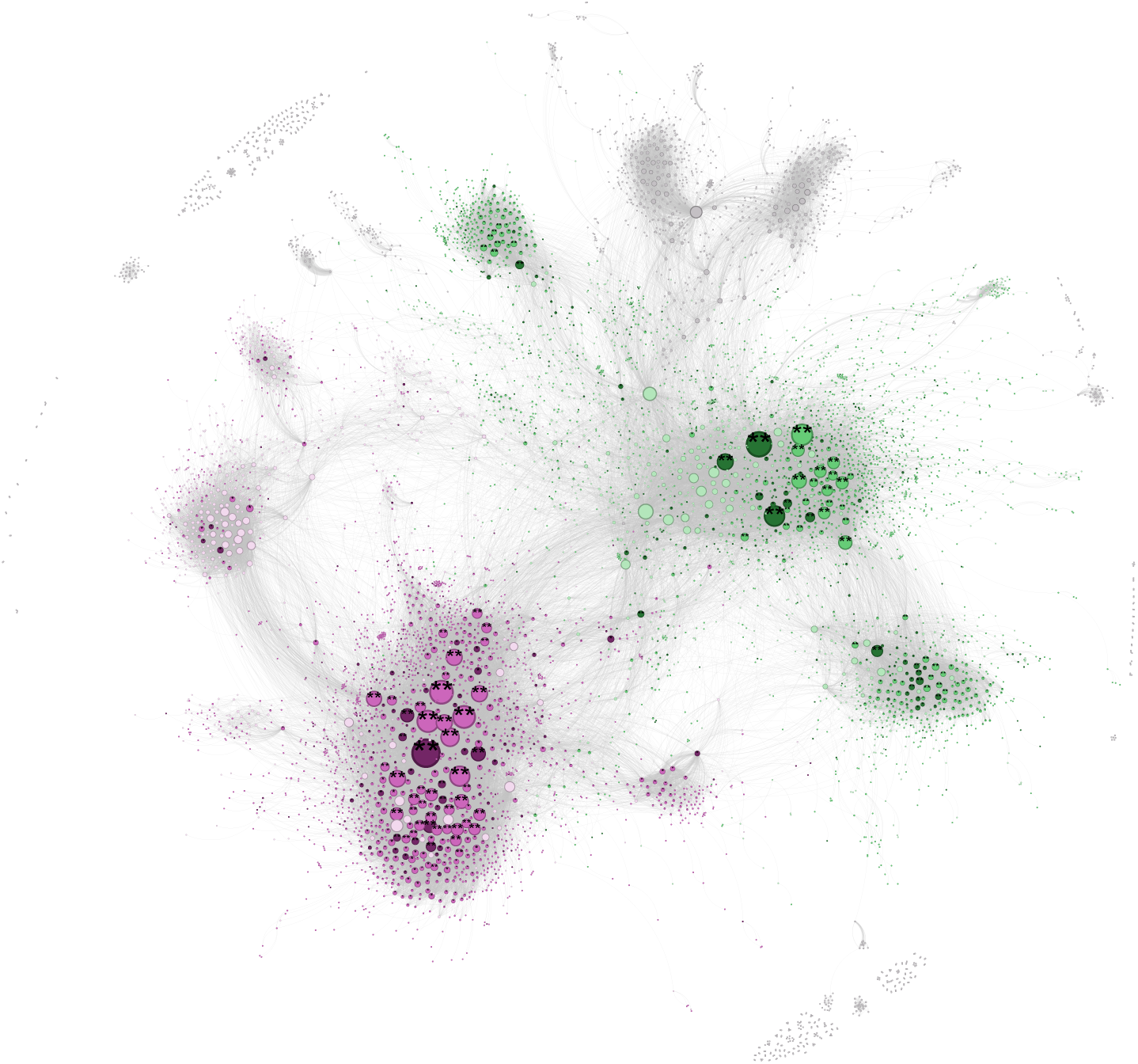
Node size corresponds to *degree centrality*. As in Figure 1, users are represented as circles. Edges connect users that were retweeted at least twice by at least ten of the same users. All edges are gray. Nodes in the two largest communities detected using the Infomap algorithm are colored in pink (anti-vaccine) and green (pro-vaccine). Shades indicate account type: non-individual and non-English accounts excluded from analyses (light); individual perceived non-expert (medium); and perceived experts (dark). Nodes outside of the two largest communities are gray. Users that rank in the top 500 and top 50 users by degree centrality within the anti-vaccine or pro-vaccine community are labeled with one and and two stars.

**Figure 14:**
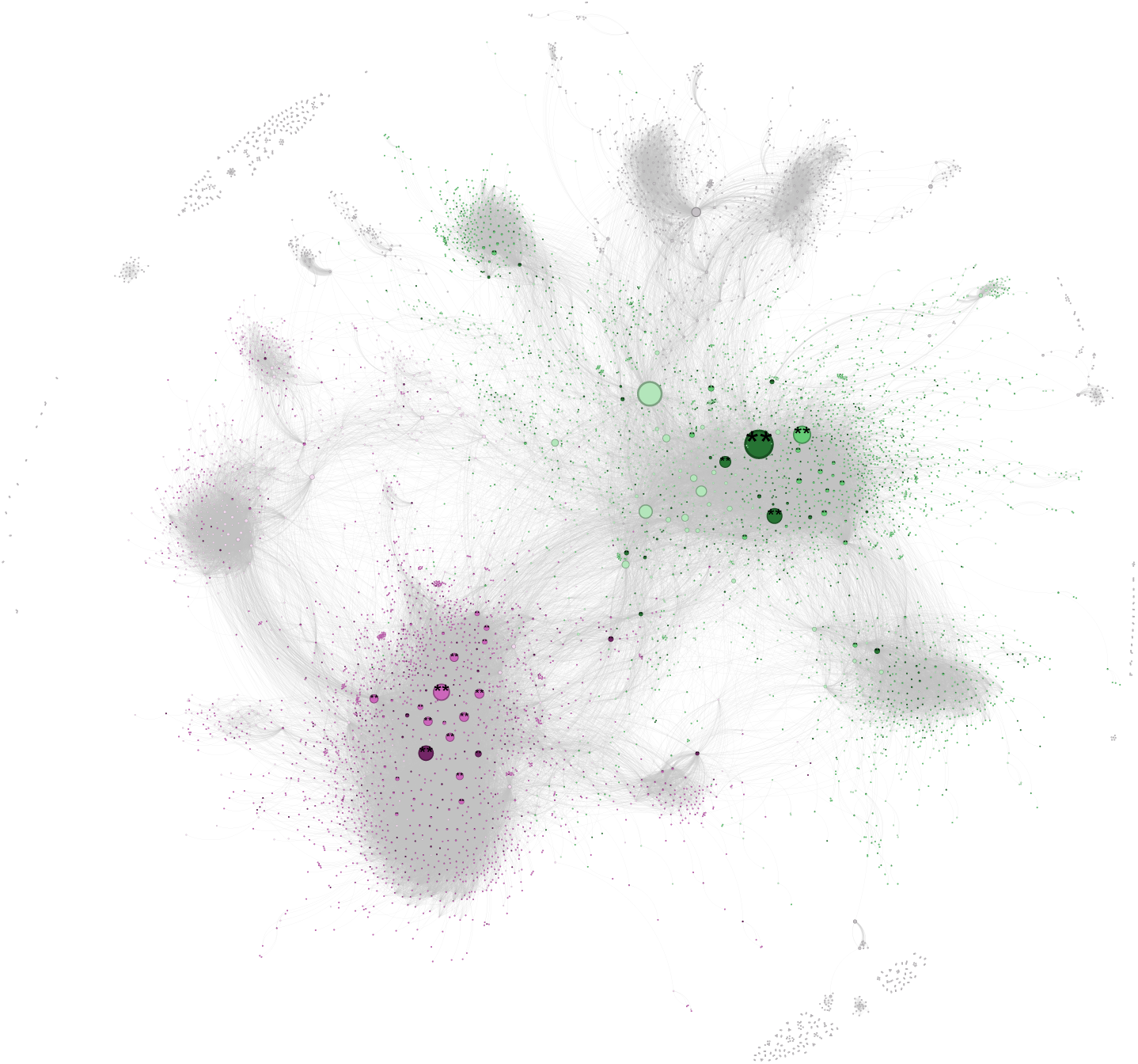
Node size corresponds to *betweenness centrality*. As in Figure 1, users are represented as circles. Edges connect users that were retweeted at least twice by at least ten of the same users. All edges are gray. Nodes in the two largest communities detected using the Infomap algorithm are colored in pink (anti-vaccine) and green (pro-vaccine). Shades indicate account type: non-individual and non-English accounts excluded from analyses (light); individual perceived non-expert (medium); and perceived experts (dark). Nodes outside of the two largest communities are gray. Users that rank in the top 500 and top 50 users by betweenness centrality within the anti-vaccine or pro-vaccine community are labeled with one and and two stars.

**Figure 15:**
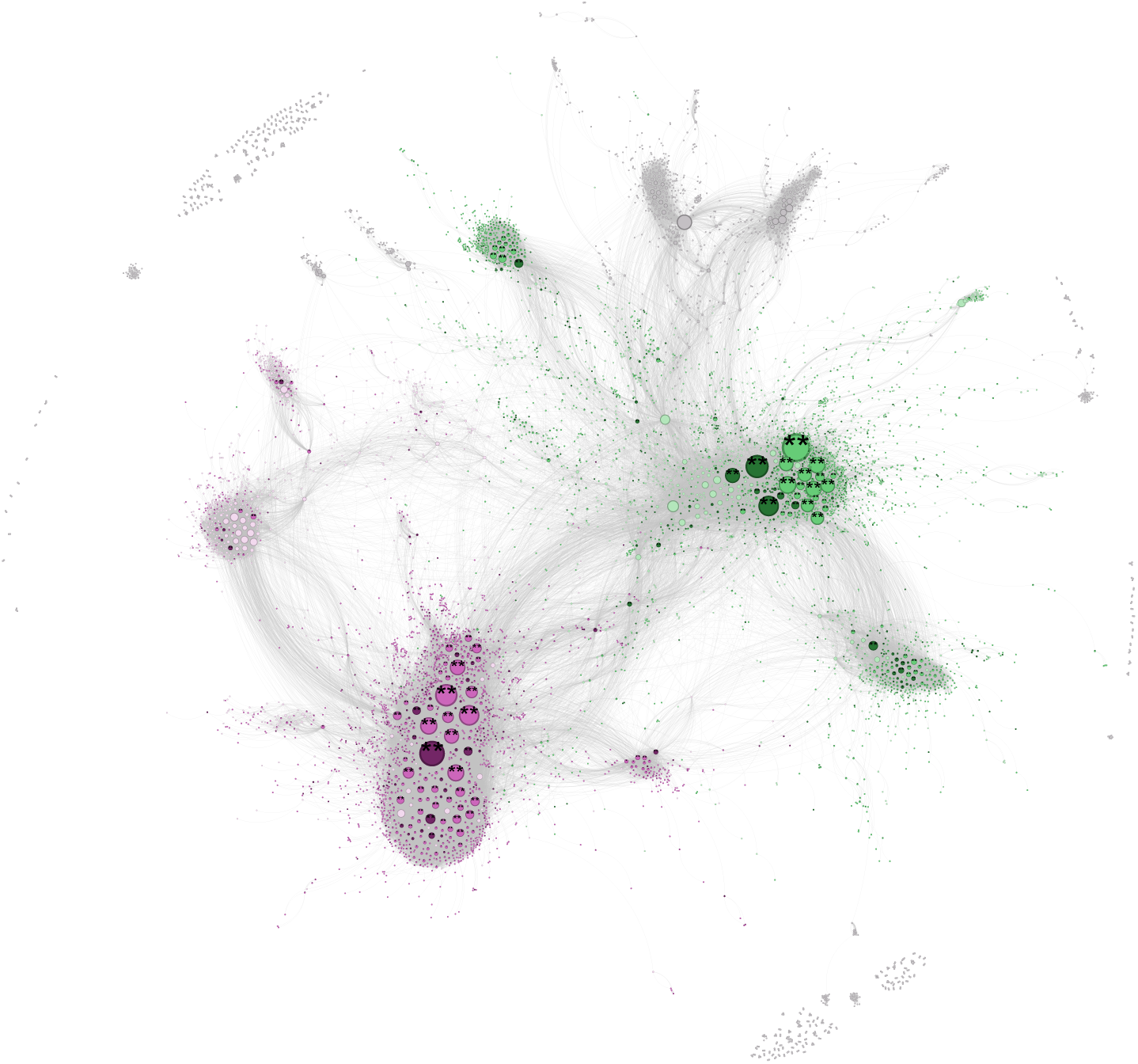
Node size corresponds to *eigenvector centrality*. As in Figure 1, users are represented as circles. Edges connect users that were retweeted at least twice by at least ten of the same users. All edges are gray. Nodes in the two largest communities detected using the Infomap algorithm are colored in pink (anti-vaccine) and green (pro-vaccine). Shades indicate account type: non-individual and non-English accounts excluded from analyses (light); individual perceived non-expert (medium); and perceived experts (dark). Nodes outside of the two largest communities are gray. Users that rank in the top 500 and top 50 users by eigenvector centrality within the anti-vaccine or pro-vaccine community are labeled with one and and two stars.

#### 7.7 Main matching analyses

This section provides sample sizes (Supplemental Table 5), covariate balance before and after matching (Supplemental Figure 16 for H1, Supplemental Figure 17 for H2, Supplemental Figure 18 for H3), and statistics related to estimated average treatment effect on the treated (ATT) (Supplemental Table 6 for H1, Supplemental Table 7 for H2, Supplemental Table 8 and Supplemental Table 9 for H3). We also visualize the estimated ATT within the anti- and pro-vaccine communities (Supplemental Figure 19) and the difference in the estimated ATT between the anti- and pro-vaccine communities (Supplemental Figure 20).

Note that Supplemental Table 8 gives that computed influence boost for perceived experts in the anti- and pro-vaccine communities and Supplemental Table 9 gives the *difference* in influence boosts for perceived experts between the anti- and pro-vaccine communities. ATTs given in Supplemental Table 8 and Supplemental Figure 5 are comparable to those in Supplemental Figure 16, Figure 17, and Supplemental Figure 19 except subcommunity was dropped as a matching covariate to enable comparison between communities.

**Table 5:**
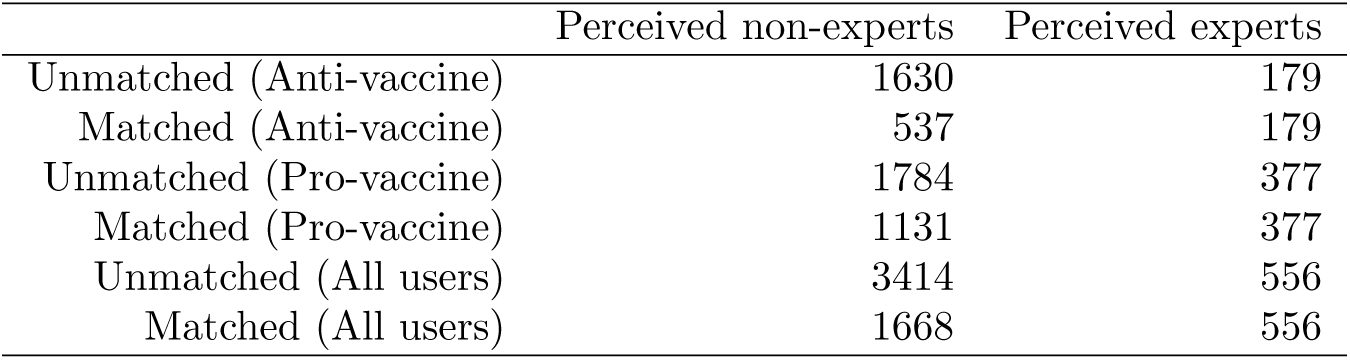
Sample size of perceived non-experts and perceived experts before and after propensity score matching in the anti-vaccine community (H1), pro-vaccine community (H2), or both (H3).

|  | Perceived non-experts | Perceived experts |
| --- | --- | --- |
| Unmatched (Anti-vaccine) | 1630 | 179 |
| Matched (Anti-vaccine) | 537 | 179 |
| Unmatched (Pro-vaccine) | 1784 | 377 |
| Matched (Pro-vaccine) | 1131 | 377 |
| Unmatched (All users) | 3414 | 556 |
| Matched (All users) | 1668 | 556 |

**Figure 16:**
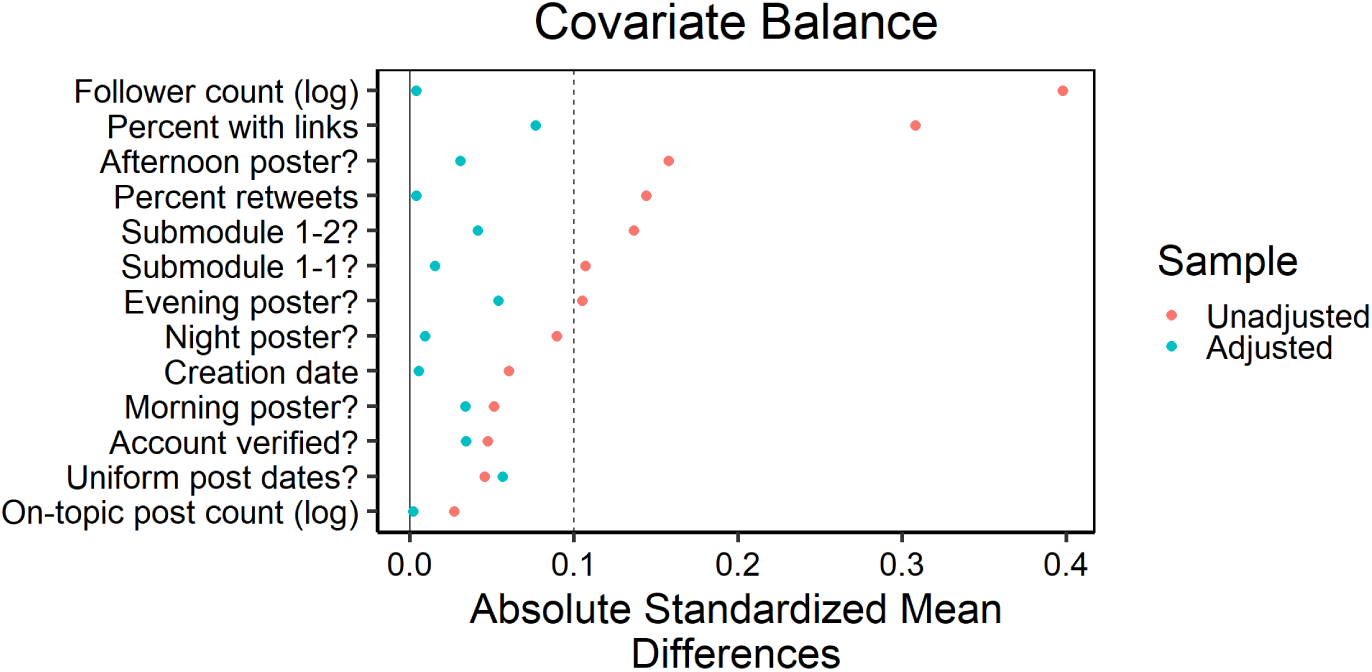
Love plot demonstrating the balance across matching co-variates for individuals in the anti-vaccine community before (orange) and after (blue) propensity score matching was performed to test H1. Each row corresponds to a different matching covariate (described in Supplement Table 2. The x-axis is absolute standardized mean difference, where values closer to zero correspond to better balance. The horizontal line indicates 0.1, the threshold below which balance is generally considered good.

**Figure 17:**
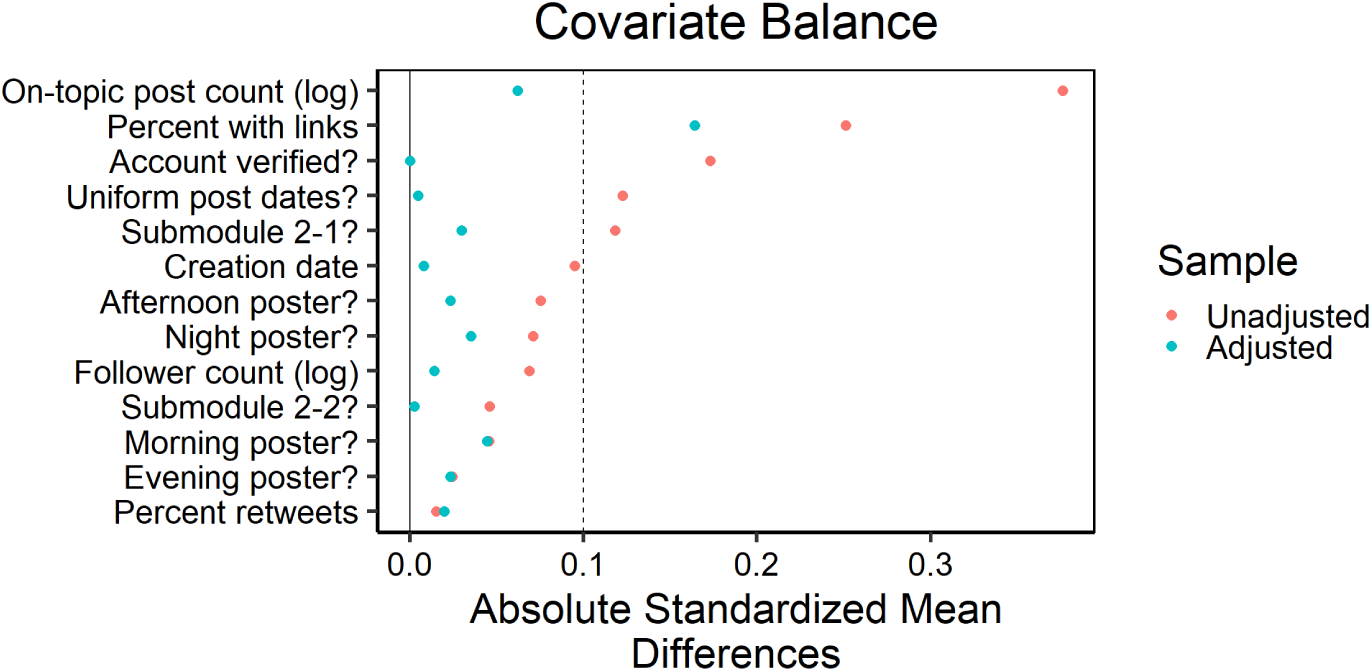
Love plot demonstrating the balance across matching covariates for individuals in the pro-vaccine community before (orange) and after (blue) propensity score matching was performed to test H2. Each row corresponds to a different matching covariate (described in Supplement Table 2. The x-axis is absolute standardized mean difference, where values closer to zero correspond to better balance. The horizontal line indicates 0.1, the threshold below which balance is generally considered good.

**Figure 18:**
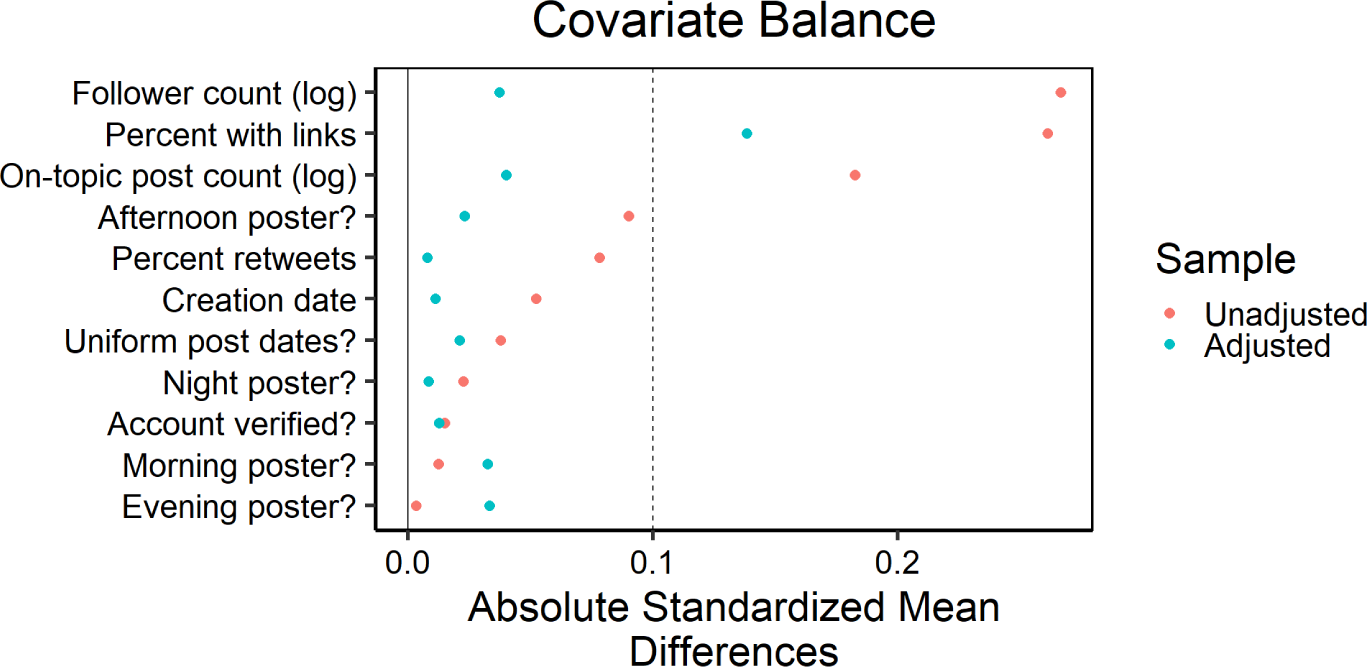
Love plot demonstrating the balance across matching covariates for individuals in the pro- and anti-vaccine communities before (orange) and after (blue) propensity score matching was performed to test H3. Each row corresponds to a different matching covariate (described in Supplement Table 2. The x-axis is absolute standardized mean difference, where values closer to zero correspond to better balance. The horizontal line indicates 0.1, the threshold below which balance is generally considered good.

**Table 6:** Average treatment effect on the treated within the anti-vaccine community (H1). Each row corresponds to results of the matching analysis for a different outcome variable, named in the first column and described in Supplemental Table 2. The estimate column gives the average treatment effect on the treated with the corresponding 95% confidence interval in parentheses. Positive values indicate greater influence for perceived experts compared to perceived non-experts. The p-value and standard error for each estimate are also provided.

| Outcome | Estimate | p-value | Std. Error |
| --- | --- | --- | --- |
| Median log(Likes) | 0.29 (0.09, 0.49) | 0.004 | 0.10 |
| h-index log(Likes) | 3.43 (1.11, 5.76) | 0.004 | 1.19 |
| Median log(Retweets) | 0.19 (0.03, 0.36) | 0.021 | 0.08 |
| h-index log(Retweets) | 1.90 (0.36, 3.44) | 0.016 | 0.78 |
| Degree Centrality | 4.96 (-6.96, 16.87) | 0.415 | 6.08 |
| Eigenvector Centrality | 3.23e-05 (-4.91e-05, 1.14e-04) | 0.437 | 0.00 |
| Betweenness Centrality | 2681.01 (-8098.77, 13460.80) | 0.626 | 5499.99 |

**Table 7:** Average treatment effect on the treated within the pros-vaccine community (H2). Each row corresponds to results of the matching analysis for a different outcome variable, named in the first column and described in Supplemental Table 2. The estimate column gives the average treatment effect on the treated with the corresponding 95% confidence interval in parentheses. Positive values indicate greater influence for perceived experts compared to perceived non-experts. The p-value and standard error for each estimate are also provided.

| Outcome | Estimate | p-value | Std. Error |
| --- | --- | --- | --- |
| Median log(Likes) | 0.36 (0.22, 0.49) | 0.000 | 0.07 |
| h-index log(Likes) | 4.33 (2.76, 5.89) | 0.000 | 0.80 |
| Median log(Retweets) | 0.13 (0.03, 0.24) | 0.012 | 0.05 |
| h-index log(Retweets) | 2.20 (1.27, 3.14) | 0.000 | 0.48 |
| Degree Centrality | 4.80 (-0.13, 9.74) | 0.056 | 2.52 |
| Eigenvector Centrality | 4.55e-05 (-2.03e-06, 9.30e-05) | 0.061 | 0.00 |
| Betweenness Centrality | 12845.45 (4429.92, 21260.98) | 0.003 | 4293.72 |

**Figure 19:**
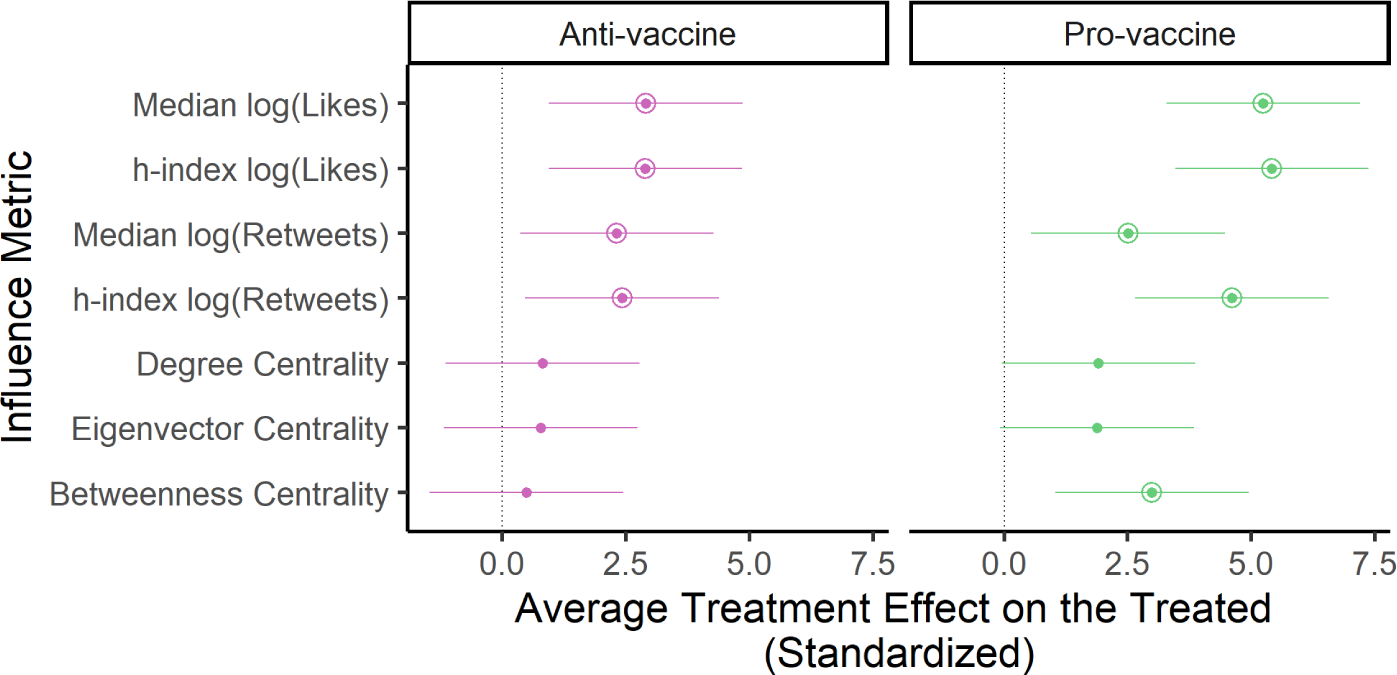
In the anti-vaccine and pro-vaccine communities, perceived experts receive greater engagements compared to other users. For each influence metric (y-axis, Supplemental Table 3, we plot the standardized average treatment effect on the treated (ATT) as a point and corresponding 95% confidence interval. Values are shown for the matching analyses comparing perceived experts and perceived non-experts within the anti-vaccine (pink, left) and pro-vaccine (green, right) communities (corresponding to Supplemental Table 6 and Supplemental Table 7 respectively) with subcommunity matching. Positive values (to the right of the vertical line) indicate an influence boost for perceived experts. Instances where the effects were significantly greater than zero (*p <* 0.05) are indicated with an additional circle around the point estimate.

**Table 8:** Average treatment effect on the treated within the anti- and pro-vaccine communities (H3). Each row corresponds to results of the matching analysis for a different outcome variable (named in the first column and described in Supplemental Table 2) and community (named in the second column). The estimate column gives the average treatment effect on the treated with the corresponding 95% confidence interval in parentheses. Positive values indicate greater influence for perceived experts compared to perceived non-experts. The p-value and standard error for each estimate are also provided.

| Outcome | Community | Estimate | p-value | Std. Error |
| --- | --- | --- | --- | --- |
| Median log(Likes) | Anti-vaccine | 0.35 (0.16, 0.55) | 0.000 | 0.10 |
| h-index log(Likes) | Anti-vaccine | 3.90 (1.59, 6.21) | 0.001 | 1.18 |
| Median log(Retweets) | Anti-vaccine | 0.25 (0.09, 0.40) | 0.002 | 0.08 |
| h-index log(Retweets) | Anti-vaccine | 2.30 (0.88, 3.72) | 0.001 | 0.73 |
| Degree Centrality | Anti-vaccine | 7.72 (-1.55, 16.98) | 0.102 | 4.73 |
| Eigenvector Centrality | Anti-vaccine | 4.32e-05 (-3.37e-05, 1.20e-04) | 0.271 | 0.00 |
| Betweenness Centrality | Anti-vaccine | 2956.88 (-9417.67, 15331.42) | 0.640 | 6313.66 |
| Median log(Likes) | Pro-vaccine | 0.38 (0.25, 0.52) | 0.000 | 0.07 |
| h-index log(Likes) | Pro-vaccine | 4.27 (2.67, 5.87) | 0.000 | 0.82 |
| Median log(Retweets) | Pro-vaccine | 0.15 (0.04, 0.25) | 0.006 | 0.05 |
| h-index log(Retweets) | Pro-vaccine | 2.20 (1.21, 3.19) | 0.000 | 0.50 |
| Degree Centrality | Pro-vaccine | 4.84 (-1.59, 11.27) | 0.140 | 3.28 |
| Eigenvector Centrality | Pro-vaccine | 4.46e-05 (-8.73e-06, 9.80e-05) | 0.101 | 0.00 |
| Betweenness Centrality | Pro-vaccine | 12874.05 (4283.48, 21464.61) | 0.003 | 4383.02 |

**Table 9:** Difference in average treatment effect on the treated between the pro-vaccine and pro-vaccine community (H3). Each row corresponds to results of the matching analysis for a different outcome variable, named in the first column and described in Supplemental Table 2. The estimate column gives the difference between the average treatment effect on the treated for the anti-vaccine community versus the pro-vaccine community with the corresponding 95% confidence interval in parentheses. Positive values indicate greater a influence boost for perceived experts in the anti-vaccine community compared to the pro-vaccine community. The p-value and standard error for each estimate are also provided.

| Outcome | Estimate | p-value | Std. Error |
| --- | --- | --- | --- |
| Median log(Likes) | -0.03 (-0.26, 0.20) | 0.804 | 0.12 |
| h-index log(Likes) | -0.37 (-3.67, 2.93) | 0.826 | 1.68 |
| Median log(Retweets) | 0.10 (-0.09, 0.29) | 0.319 | 0.10 |
| h-index log(Retweets) | 0.10 (-1.95, 2.15) | 0.922 | 1.05 |
| Degree Centrality | 2.88 (-12.19, 17.95) | 0.708 | 7.69 |
| Eigenvector Centrality | -1.47e-06 (-1.12e-04, 1.09e-04) | 0.979 | 0.00 |
| Betweenness Centrality | -9917.17 (-28717.04, 8882.70) | 0.301 | 9591.95 |

**Figure 20:**
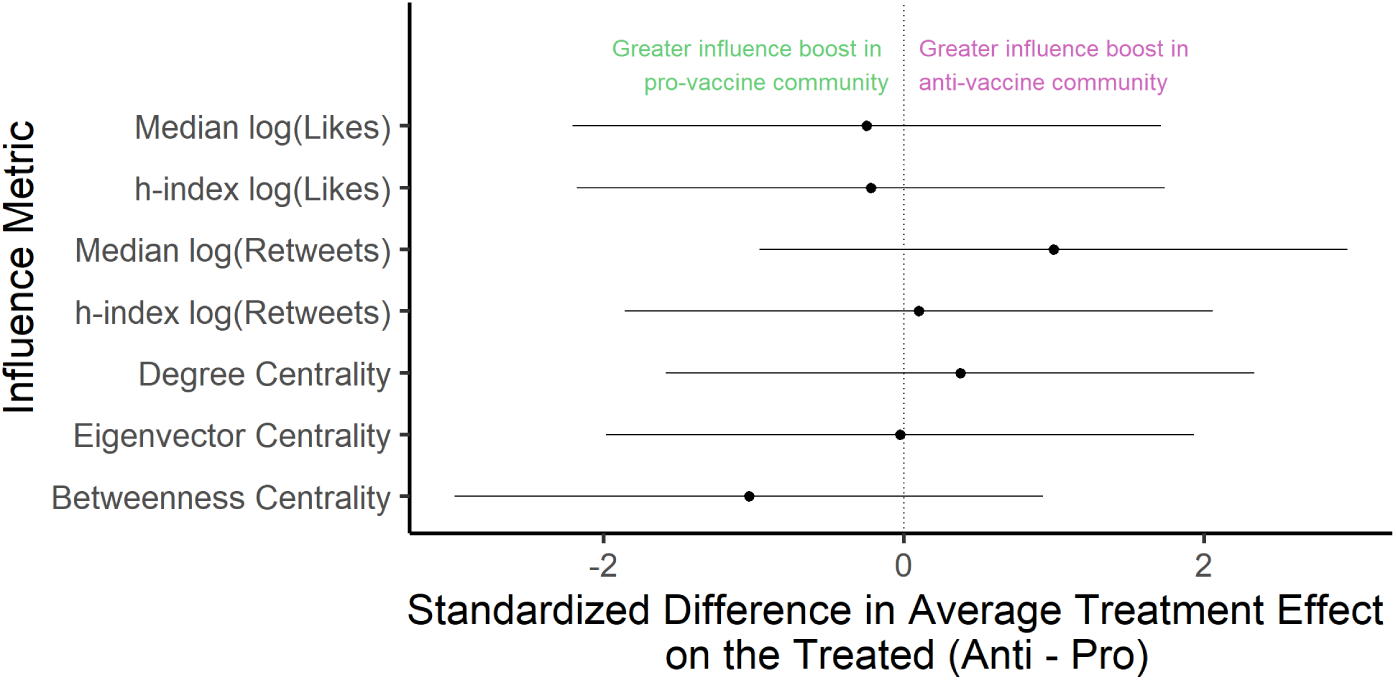
There is no significant difference in the influence boost for perceived experts between the anti- and pro-vaccine communities. For each influence metric (y-axis, Supplemental Table 3, we plot the difference in the standardized average treatment effect on the treated (ATT) between the pro- and anti-vaccine communities as a point and corresponding 95% confidence interval (Supplemental Table 9). Positive values (to the right of the vertical line) indicate a greater influence boost for perceived experts in the anti-vaccine commmunity compared to the pro-vaccine community. None of the differences were significant at the *p <* 0.05 level.

#### 7.8 Distribution of centrality values

This section further investigates why, despite being overrepresented amongst users with the greatest centrality Figure 3, perceived experts do not, on average, have greater centrality than the matched set of perceived non-experts Figure 5. Matching generally reduces differences the frequency distribution for perceived experts versus perceived non-experts across centrality metrics, suggesting that matching covariates may contribute to the greater frequency of perceived experts with high centrality. Balancing therefore may partially reduce the difference in mean centrality between perceived experts and perceived non-experts. However, perceived experts generally remain overrepresented in the right tail of the distribution, even while this effect does not lead to a significant increase in the mean centrality of perceived experts.

**Figure 21:**
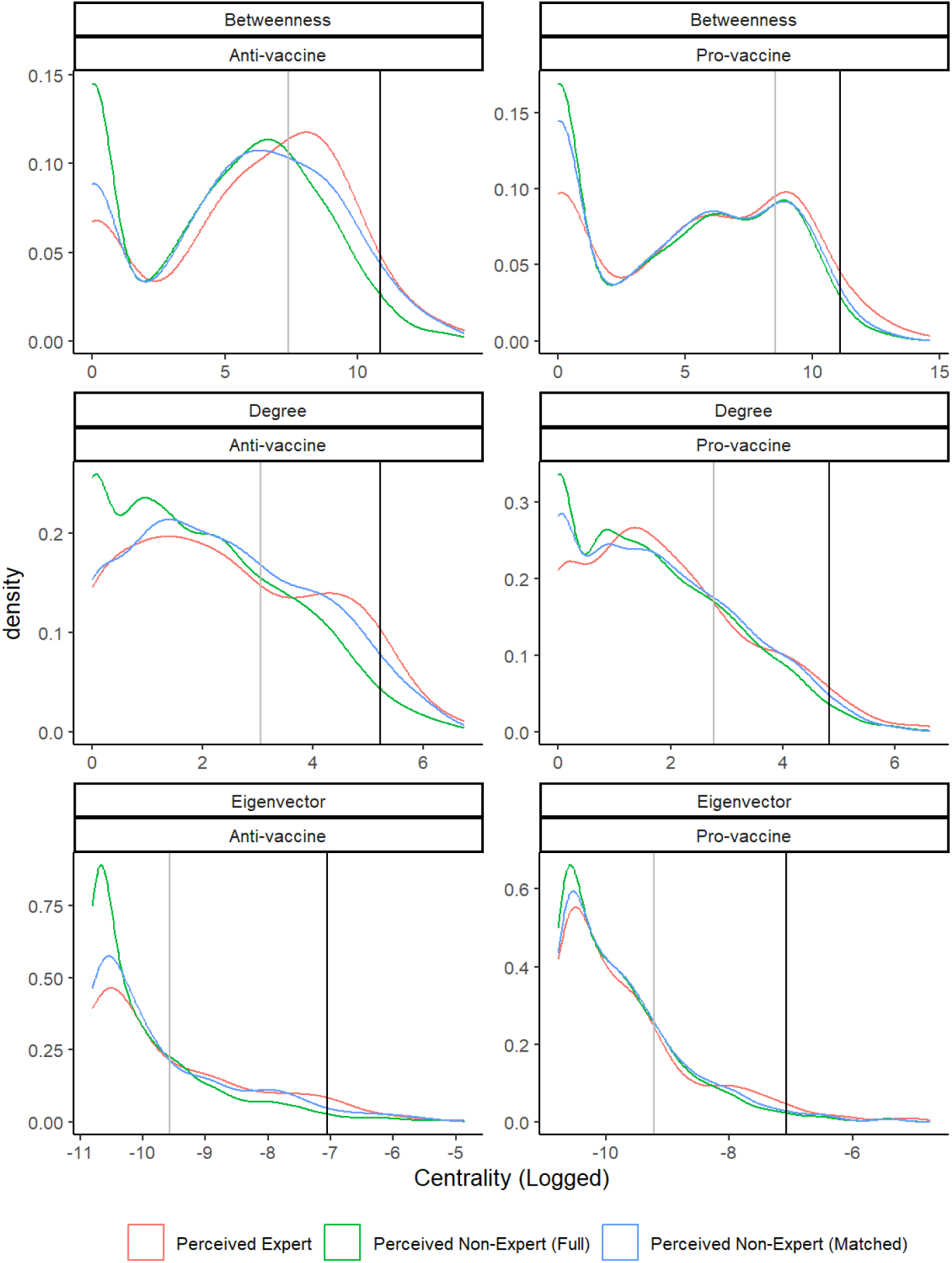
Matching reduces the proportion of perceived non-experts with low centrality. Each plot shows the frequency distribution of users for a given logged centrality metric. Values are log-transformed because distributions are highly skewed. Lines are colored by the subset of users: perceived experts (red), perceived non-experts in the full community prior to matching (green), and perceived non-experts in the community subset for matching (blue). Values to the right of the vertical lines indicate users in the top 500 (gray) or 50 (black) users ranked by the given centr_5_a_4_lity metric. Each row corresponds to a different centrality metric: betweeenness (top), degree (middle), and eigenvector (bottom). Each columns corresponds to a different community: anti-vaccine (left) or pro-vaccine (right).

#### 7.9 Sensitivity to matching specifications

This section shows that propensity score matching results are generally robust to k, the number of nearest neighbors to which each perceived expert is matched (Supplemental Figure 22), and the exclusion of any one matching covariates (Supplemental Figure 23 for balance, Supplemental Figure 24 for ATT). We do note that the significant ATT for betweenness centrality in the pro-vaccine community does not hold if a variable other than percent with links is excluded, suggesting that this result in particular may be sensitive to matching on this covariate. Poor balance is generally achieved for the percent with links covariate.

**Figure 22:**
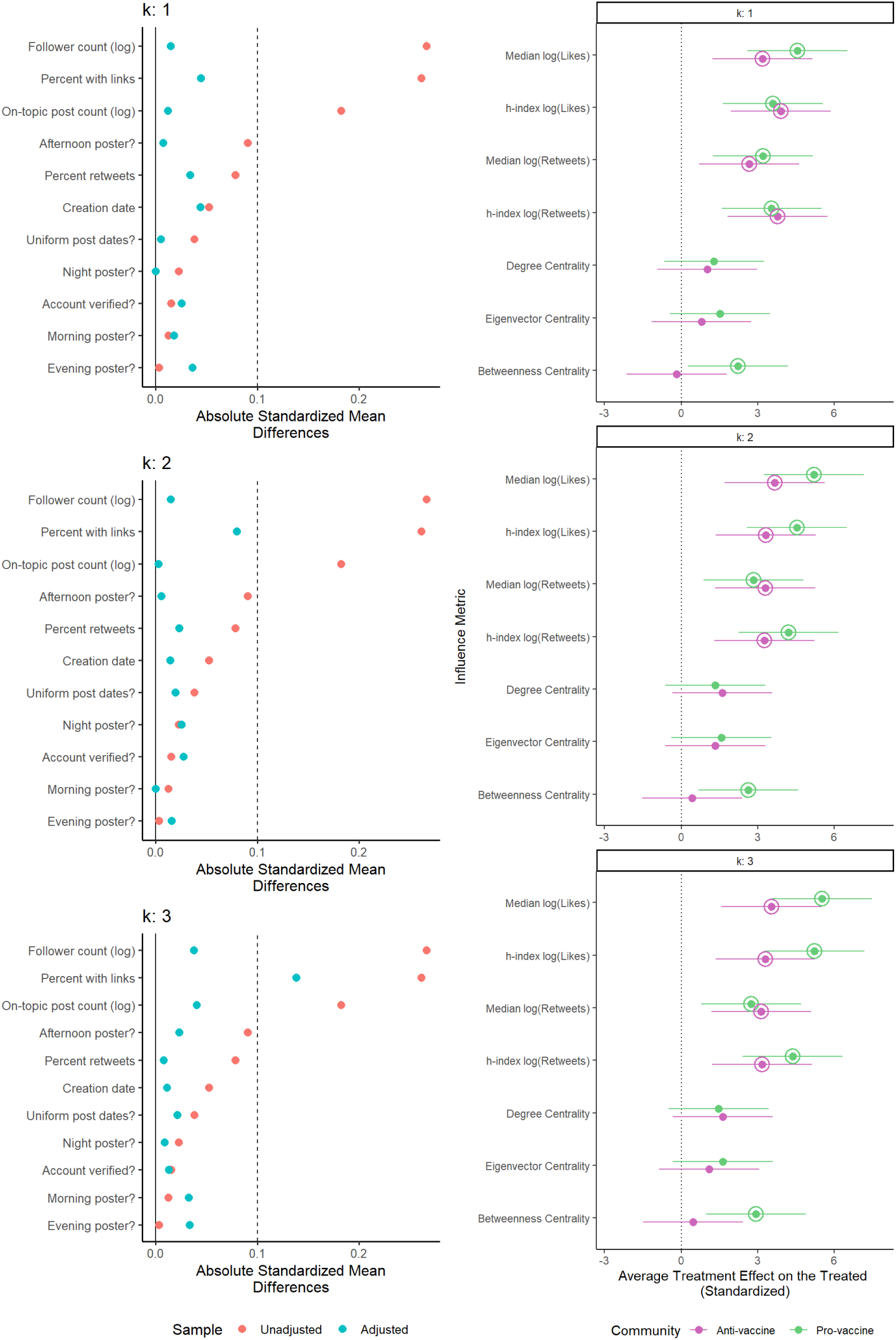
Matching balance and results are robust to k, the number of nearest neighbors to which each perceived expert is matched. Caption continued on following page. Each row corresponds to a different value of k: 1 (top), 2 (middle), or 3 (bottom). The left column contains Love plots demonstrating the balance across matching covariates for individuals in the pro- and antivaccine communities before (orange) and after (blue) propensity score matching was performed. Within a plot, each row corresponds to a different matching covariate (described in Supplement Table 2. The x-axis is absolute standardized mean difference, where values closer to zero correspond to better balance. The horizontal line indicates 0.1, the threshold below which balance is generally considered good. The right column shows the standardized average treatment effect on the treated (point) and corresponding 95 % confidence interval for each influence metric (y-axis, Supplemental Table 3). Values are shown for the third matching analysis directly comparing the anti-vaccine (pink) and pro-vaccine (green) communities. Positive values (to the right of the vertical line) indicate an influence boost for perceived experts. Instances where the average treatment effect on the treated was significantly greater than zero (*p <* 0.05) are indicated with an additional circle around the point estimate.

**Figure 23:**
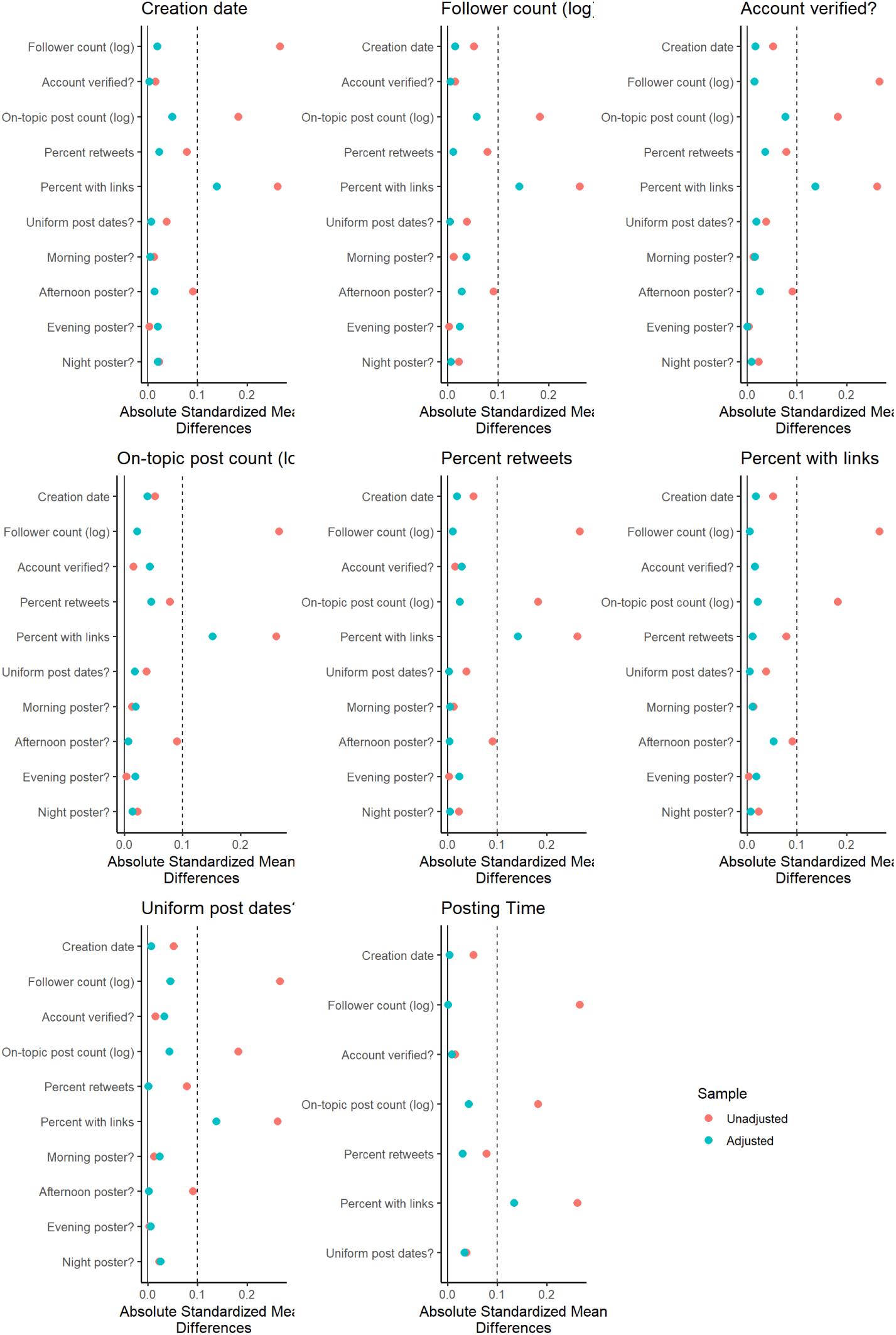
Matching balance is robust to matching covariates. Matching covariates were dropped one at a time to detect whether inclusion of a single covariate affected balance across other_5_m_8_ atching covariates. Caption continued on following page. Each plot corresponds to a different dropped covariate indicated in the panel title (described in Supplement Table 2). Note that the variable *posting time* encompasses the three binary variables: *morning poster*, *evening poster*, and *night poster*. Each Love plots demonstrates the balance across matching covariates for individuals in the pro- and anti-vaccine communities before (orange) and after (blue) propensity score matching was performed. Each row within a figure corresponds to a different matching covariate. The x-axis is absolute standardized mean difference, where values closer to zero correspond to better balance. The horizontal line indicates 0.1, the threshold below which balance is generally considered good.

**Figure 24:**
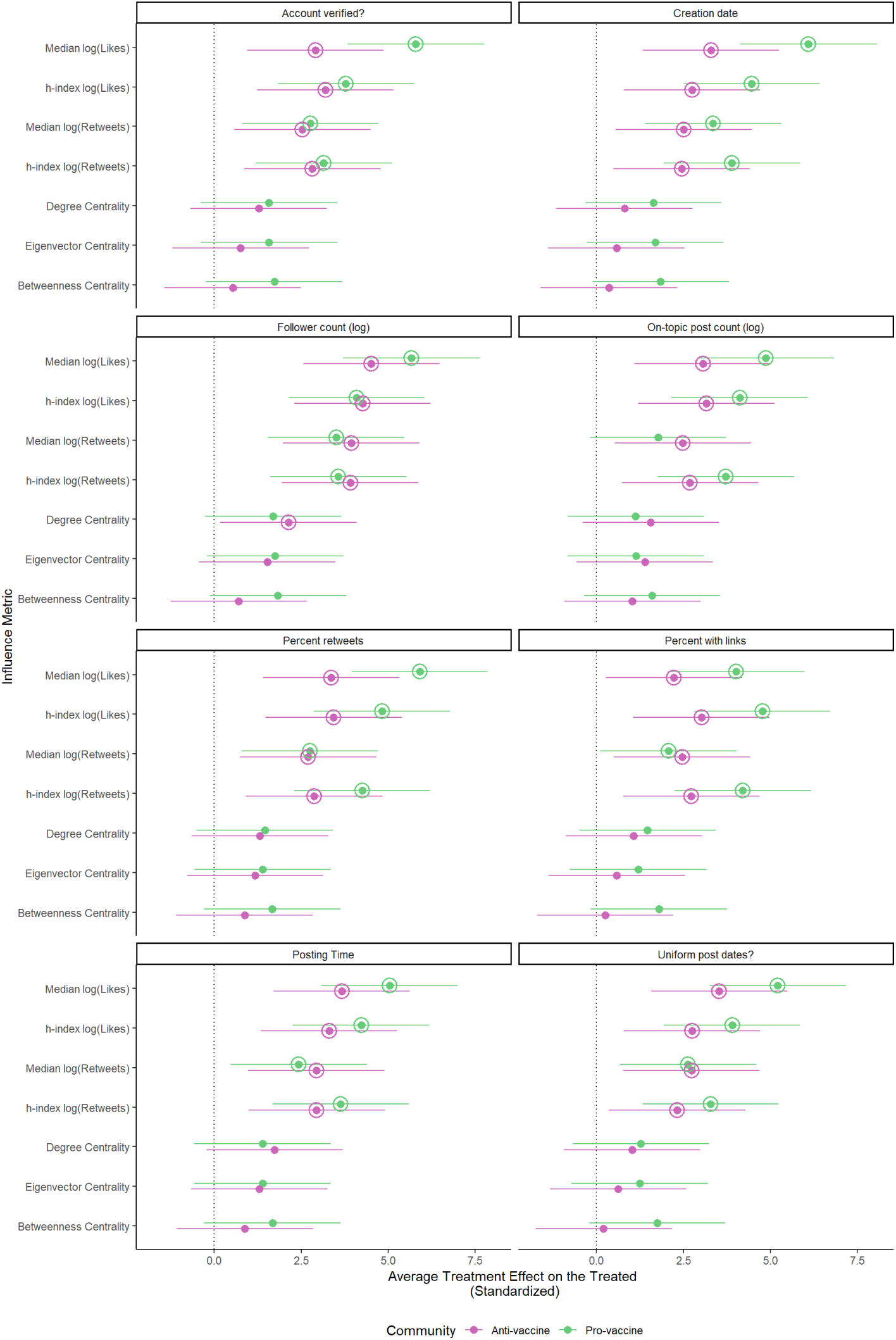
Average treatment effect on the treated (ATT) is robust to matching covariates. Matching covariates were dropped one at a time to detect whether inclusion of a single cov_6_a_0_riate affected ATT. Caption continued on following page. Each plot corresponds to a different influence metric indicated in the panel title (described in Supplemental Table 3). Plots show the standardized average treatment effect on the treated (point) and corresponding 95 % confidence interval depending on which matching covariate was dropped (Supplement Table 2). Note that the variable *posting time*encompasses the three binary variables: *morning poster*, *evening poster*, and *night poster*. Values are shown for the third matching analysis directly comparing the anti-vaccine (pink) and pro-vaccine (green) communities. Positive values (to the right of the vertical line) indicate an influence boost for perceived experts compared to other individual users. Instances where the average treatment effect on the treated was significantly greater than zero (*p <* 0.05) are indicated with an additional circle around the point estimate.

## 8 Works Cited

